# Abnormalities in cortical pattern of coherence in interictal migraine detected using ultra high-density EEG

**DOI:** 10.1101/2020.07.17.20156299

**Authors:** Alireza Chamanzar, Sarah M Haigh, Pulkit Grover, Marlene Behrmann

**Author notes:** Co-first authors (Alireza Chamanzar, Sarah M Haigh).

## Abstract

Individuals with migraine generally experience photophobia and/or phonophobia during and between migraine attacks. Many different mechanisms have been postulated to explain these migraine phenomena including abnormal patterns of connectivity across the cortex. The results, however, remain contradictory and there is no clear consensus on the nature of the cortical abnormalities in migraine. Here, we uncover alterations in cortical patterns of coherence (connectivity) in interictal migraineurs during the presentation of visual and auditory stimuli, as well as at rest, to capitalize on the sensory sensitivities that are characteristic of migraine, and to reconcile the conflicting literature on migraine cortical connectivity. We used a high-density electroencephalography (HD-EEG) system, with 128 customized electrode locations, to measure inter- and intra-hemispheric coherence from 17 individuals with migraine (12 female) in the interictal period, and 18 age- and gender-matched healthy control subjects, during visual (vertical grating pattern) and auditory (modulated tone) stimulation which varied in temporal frequency (4 and 6Hz), and during rest. To ensure that participants were attending, participants performed a color detection task at fixation. Analyses of the EEG signal included characterizing the inter- and intra-hemisphere coherence between the scalp EEG channels over 2-second time intervals and over different frequency bands at different spatial distances and spatial clusters, and Pearson’s correlation coefficients (PCCs) were estimated at zero-lag. Repeated measures (between-group) analyses-of-variance with post hoc multiple comparison correction were conducted. Relative to controls, migraineurs exhibited significantly (i) faster color detection performance; (ii) lower long-distance spatial coherence of alpha-band activity during both evoked conditions, regardless of the stimulation frequency; (iii) lower coherence between the right frontal cluster and all clusters in the left hemisphere (inter-hemisphere coherence) during 4Hz auditory and visual stimulation; and (iv) lower long-distance coherence (in all frequency bands) between the right occipito-parietal cluster and all other clusters during the (4Hz and 6Hz) visual stimuli. No significant group differences were observed in the resting state data. The abnormal patterns of EEG coherence in interictal migraineurs during visual and auditory stimuli may be associated with cortical hyper-excitability in migraineurs.

## Introduction

More than 38 million people in the United States suffer from migraine (Migraine Research Foundation (MRF), 2020), a neurovascular condition which manifests as episodes of headache, accompanied by other autonomic and neurological symptoms. The incidence of migraine worldwide is high, with current estimates of about 10% of people being affected (National Institute of Neurological Disorders and Stroke (NINDS), 2019). Neurophysiological, morphometric, and functional imaging studies have focused on uncovering the pathogenesis, biomarkers, and effective treatments for this sometimes debilitating disorder. Notwithstanding this large scientific effort, there is still no consensus on the underlying mechanism/s that give rise to migraines.

One candidate mechanism for migraine is altered functional cortical connectivity. The majority of functional connectivity measures are from fMRI resting-state (Mainero et al., 2011; Russo et al., 2012; Hadjikhani et al., 2013; Tessitore et al., 2015; Tedeschi et al., 2016; Soheili-Nezhad et al., 2019) and EEG scans (Cao et al., 2016; Wu et al., 2016; for a review, see De Tommaso, 2019). Connectivity alterations have been identified in multiple cortical regions in the pre-ictal and ictal migraine phases (Chong et al., 2017), suggesting that abnormal functional connectivity is not a consequence of the migraine attack itself. However, reviews of the functional connectivity findings from resting scans in migraine show inconsistencies across studies (Skorobogatykh et al., 2019). Some of the more consistent findings are abnormal connectivity between brain areas associated with pain, and between sensory areas (Hodkinson et al., 2016a), and strength of connectivity was related to headache severity (for a review, see Schwedt et al., 2015).

Another potential mechanism, that may be related to the altered functional connectivity (Hodkinson et al., 2016b), is greater cortical hyper-excitability (Welch et al., 1990; Aurora and Wilkinson, 2007; Tfelt-Hanson and Koehler, 2011). The majority of evidence for cortical hyper-excitability in migraine originates from studies on sensory processing due to the symptoms of photophobia and phonophobia that are common in migraine (ICHD 2018). Migraine and individuals with photosensitive epilepsy show similar visual sensitivities (Wilkins et al., 1984; Haigh et al., 2012) and migraineurs show heightened cortical responses to stimuli that can evoke photoparoxysmal EEG activity in patients with photosensitive epilepsy (Huang et al., 2003; Coutts et al., 2012; Haigh et al., 2019). Epilepsy is associated with a hyper-excitable cortex and so the similarities between migraine and epilepsy suggest a similar pathology.

Studies focusing on sensory-evoked responses in migraine have generally found larger responses, in both fMRI (Huang et al., 2003, 2011; Coutts et al., 2012; Cucchiara et al., 2015) and EEG (Ambrosini et al., 2017; Haigh et al., 2019), and reduced habituation to repeated stimuli (Kropp and Gerber, 1993) compared to controls. This evidence is consistent with cortical hyper-excitability in migraine. The majority of the evidence for cortical hyper-excitability in migraine focuses on the visual system to stimuli such as checkerboards, repetitive flashes or pattern reversal stimulation (Ambrosini et al., 2003; Aurora and Wilkinson, 2007). One study that is particularly pertinent for the current investigation is that larger amplitude steady-state visual evoked potentials (SSVEPs) were found in interictal migraineurs, and these stimuli harmonized oscillations of different cortical areas, including visual areas (Mehnert et al., 2019). The bias towards abnormal visual functioning may be related to the prevalence of auras which are more likely to be visual (Hadjikhani et al., 2001).

On the other hand, few studies have investigated the neural correlates of auditory processing in migraine. One study found that children with migraine exhibited deficits in tests of acoustic timing (Agessi et al., 2017), while another found that otoacoustic emissions were lower in women with migraine, but only at lower temporal frequencies (Joffily et al., 2016). These findings may be related to the abnormal electrophysiological responses to auditory stimuli. Auditory brainstem responses (ABRs) and auditory evoked potentials (AEPs) are abnormal in migraine (De Tommaso et al., 2004; Sable et al., 2004), which corresponds with the reduced grey matter in the brainstem (Marciszewski et al., 2018). Together, these observations indicate abnormalities in auditory and visual processing in migraine that may be associated with or impacted by the phonophobia and photophobia.

Therefore, in the current study, we investigated functional connectivity in migraine compared to headache-free controls during rest, as well as sensory stimulation, to attempt to reconcile the conflicting literature on migraine cortical connectivity. Owing to the heightened sensory sensitivity in migraine, abnormal functional connectivity may be easier to detect during sensory stimulation compared to rest. We chose to use EEG as its superior temporal resolution allows for connectivity to be assessed in different frequency bands. Previous studies have reported frequency band-specific differences in migraineurs compared to controls. However, the findings are mixed depending on where on the scalp the abnormalities appear, and whether the responses were recorded at rest or are stimulus-evoked. For example, relative to controls, reports suggest an increase in the delta or theta band during rest (Lia, et al., 1995; Bjork et al., 2009a; Tolner, et al., 2019), and during visual evoked responses in interictal migraineurs (Genco et al., 1994; Sand, 2003). Whereas others report low overall coherence in theta band (Frid et al., 2020), low inter-hemispheric coherence in delta, beta, and alpha bands, and high intra-hemispheric coherence (in all frequency bands) in female interictal migraineurs during rest (using low density EEG; Koeda, et al., 1999). In addition, decreased power in the alpha band (Lia, et al., 1995; Sand, 2003) or increased power asymmetry in the alpha band have also been reported during resting scans (O’Hare et al., 2018; Fachetti et al., 1990), and during SSVEP stimulation (Sand 1991; Bjork, et al., 2011; Tolner, et al., 2019). In another study, using high frequency flash stimuli, increased phase synchronization in alpha band in interictal migraine patients without aura was reported (de Tommaso et al., 2013). Decreased cortical coherence after photic stimulation, and increased coherence during the resting state in female interictal migraine with aura have also been reported in all of the frequency bands except gamma (Mendonça-de-Souza et al., 2012). Other, more recent studies, however, have reported lower EEG power and coherence in fronto-central and parietal regions (Cao et al., 2016), all in interictal migraine patients, during the resting state, and in all of the frequency bands except gamma.

There are many potential explanations for these contradictory results including differences in the method of data acquisition, for example, using different EEG densities (Koeda, et al., 1999), and differing between rest and sensory-evoked scans. The goal of the current investigation is to conduct a comprehensive analysis of cortical connectivity in individuals with migraine versus headache-free controls. To do this, we acquired data using a custom-designed EEG cap with ultra-high-density coverage (approximately 14 mm center-to-center electrode distance) over visual, parietal, and frontal regions (Haigh et al., 2019). EEG signals were obtained from participants under three conditions: visual stimulation (vertical grating patterns), auditory stimulation (modulated tone), and resting state in order to identify any unique as well as any common patterns of abnormality in the spatial coherence in migraineurs versus control participants.

Although EEG amplitude measurements or calculation of power spectra are simple to compute, we elected to study coherence, which is considered to be a reliable measure of synchronization of the electro-cortical activities (Markovska-Simoska, et al., 2018) and has been used to investigate connectivity in migraine previously (refs where applicable). Because spatial coherence drops as a function of distance between electrodes, we analyzed the estimated coherence as a function of inter-electrode distance (link length). We chose two commonly used stimulation frequencies of 4Hz and 6Hz (Akben et al., 2009; Akben et al., 2016; Chamanzar et al., 2020), as two viable frequencies for both visual and auditory stimulation to ensure any comparisons across sensory modalities and between groups were not temporal frequency-specific. If sensory stimulation primarily drives abnormal connectivity in migraine, then we predicted group differences would be evident during evoked but not rest conditions. If, however, the disturbed connectivity is independent of input, then we might see coherence differences in both the evoked and rest scans.

## Materials and methods

### Participants

Seventeen adults with migraine (mean age 27.6 years old; range 19-54 years; 12 female), and eighteen age- and gender-matched headache-free controls (mean age 27.9 years old; range 19-54 years) were recruited from Carnegie Mellon University and from the surrounding Pittsburgh area for the study. Participants were paid $50 for their participation.

All individuals with migraine satisfied the International Headache Society (IHS) criteria with 12 of them classified as having migraine with aura (ICHD-3 1.2), and five as having migraine without aura (ICHD-3 1.1). Three in the migraine-with-aura group took medication (two took Triptans and the other had Botox). We excluded these patients from all the analyses in this paper to make sure the medication was not affecting the results. In addition, we pooled together migraineurs, with and without aura, in our analyses. The limited number of migraineurs without aura (five) makes the comparisons between migraineurs with and without aura underpowered and difficult to interpret. For spatial coherence analysis to be an effective biomarker of migraine, we anticipate a medium to large effect size. For example, in (Mendonça-de-Souza et al., 2012) 11 migraine patients were used for a spatial coherence analysis. Therefore, abnormal patterns of coherence should be detectable with the 14 participants with migraine (mean age 25.9 years old; range 19-47 years; 9 female) in our study, while excluding the patients who took medication.

Control participants were classified as being headache-free if, by self-report, they had never had a headache or had infrequent headaches that were less than moderately painful and had no co-occurring sensory disturbances.

Participants had no neurological or psychological diagnoses (except for migraine), no previous severe head injury or concussion, had normal hearing, and normal or corrected-to-normal vision by self-report. All procedures were approved by the Carnegie Mellon University Institutional Review Board. Written informed consent form was received from each subject before starting each recording session.

### Stimuli

MATLAB (MathWorks) and the Psychtoolbox extension (Brainard, 1997; Pelli, 1997; Kleiner et al, 2007) were used to generate and present the stimuli.

#### Visual Stimulation

Vertical sinusoidal-wave achromatic gratings presented at 0.05 cpd, subtended 5.7 degrees of visual angle in diameter in the center of the screen at a viewing distance of 1 meter (see Fig. 1). Gratings were filtered using a spatial 2D Gaussian filter. The gratings alternated contrast at 4Hz or at 6Hz for 2s followed by an inter-stimulus interval that varied between 1-1.5s. Each temporal frequency was presented 100 times and the order of temporal frequency was randomized. A central fixation cross (0.5 cpd) was presented throughout the duration of the experiment and was superimposed on the central grating.

**Figure 1.**
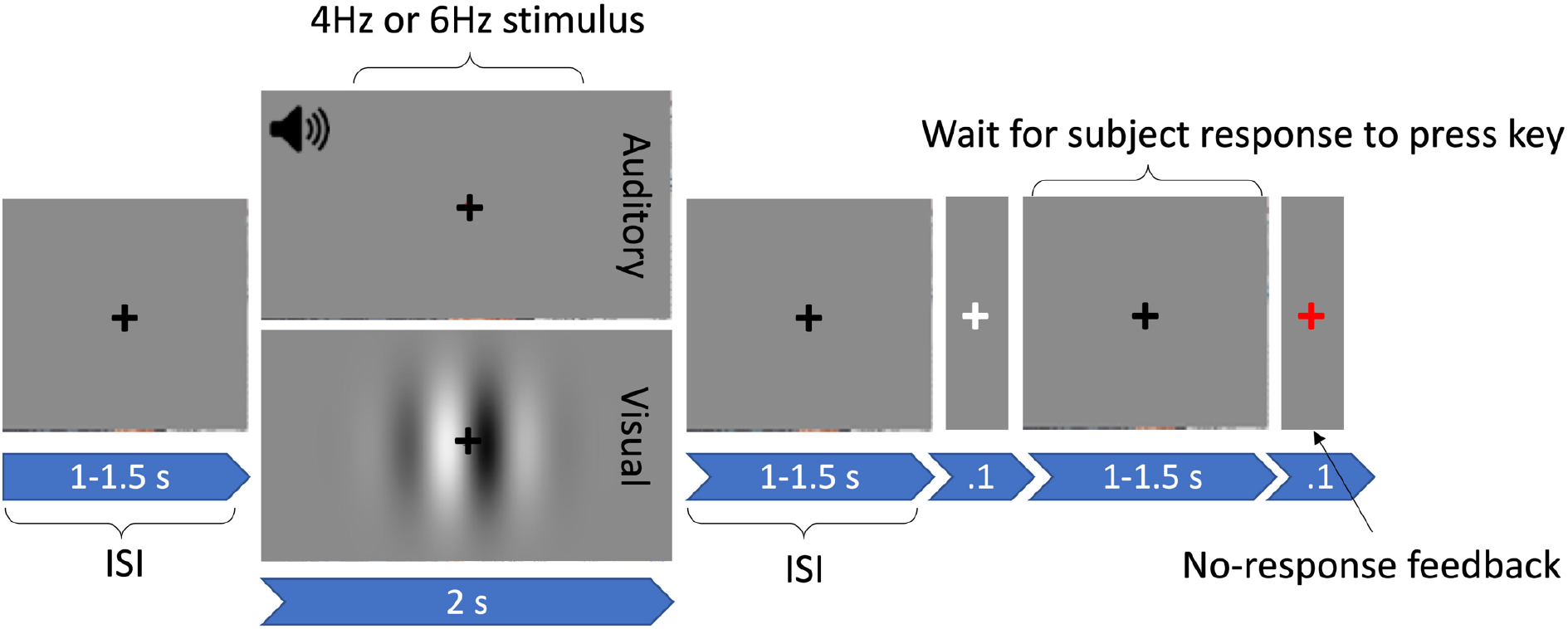
Structure of auditory and visual trials. A fixation cross appeared centrally, jittered for between 1-1.5s. A stimulus, either auditory or visual, was then presented for 2s followed by a 1-1.5s inter-stimulus-interval (ISI), consisting of a grey screen and a fixation cross. Participants pressed the spacebar whenever the cross flashed white (for 0.1s). If they did not respond, the fixation cross turned red (for

#### Auditory Stimuli

The auditory stimuli were 1kHz tones, modulated by a sinusoidal 4 or 6Hz carrier frequency and were presented for 2s. The stimuli were sampled at 44.1kHz with 16-bit resolution. Each modulator frequency was presented 100 times. Tones were separated by an inter-stimulus interval between 1-1.5s where no sound was played. Tones were presented over insert earphones (Etymotic Research Inc.). A grey screen with a black central fixation cross was presented for the duration of the experiment (see Fig. 1).

#### Resting state

The resting state condition consisted of a single black fixation cross at the center of the grey screen for the duration of the scan.

### Procedure

The resting EEG scan was always completed first followed by the stimulation conditions so that the sensory stimuli would not contaminate the resting EEG signal. During the resting scan, participants were asked to keep their eyes open and fixated on the black central cross. Six blocks of 2-minutes (12 minutes total) were recorded, with a break in between each block.

The order of the visual and auditory scans was counterbalanced across participants. For both scans, the procedure was the same. As shown in Fig. 1, stimuli were presented for 2s and preceded and followed by a grey screen with a fixation cross (inter-stimulus interval of 1-1.5s; random with uniform distribution). Participants were asked to ignore the stimuli and to attend to the fixation cross. They were required to press the spacebar whenever the cross flashed white (for 0.1s), which occurred randomly on 10% of trials. If the participant did not respond, the fixation cross turned red for 0.1s. Four blocks of 50 trials (25 of the 4Hz stimuli, 25 of the 6Hz) were presented and in each block the stimulation frequencies were randomly ordered. Each block of trials was approximately three minutes, which means that we recorded around 6 minutes of EEG data for each of the auditory and visual conditions.

### EEG Recording

A 128 channel BioSemi Active II system (BioSemi, Amsterdam, Netherlands) was used to record the EEG signals at 512Hz sampling frequency using a 24-bit A/D converter. We used a custom-designed high-density EEG nylon cap with electrodes specifically positioned so as to cover with high resolution just over central occipital, parietal, and frontal areas (the locations of alterations typically reported in previous studies of migraine). More details of the electrode locations are available in our previous paper (Haigh et al., 2019). Electrodes were placed approximately 1.4 cm distance from one another. A 2D map of the electrode locations is shown in Fig. 2. An additional seven electrodes were placed around the head. To detect the electrooculography signals, four electrodes were placed around eyes: one electrode above and one below the right eye, and one on the outer canthi of each eye. For recording of electrocardiography signals, one electrode was placed on the collar bone. Two electrodes were placed on the mastoids. Standard BioSemi CMS and DRL electrodes were used as online references for all electrodes.

**Figure 2.**
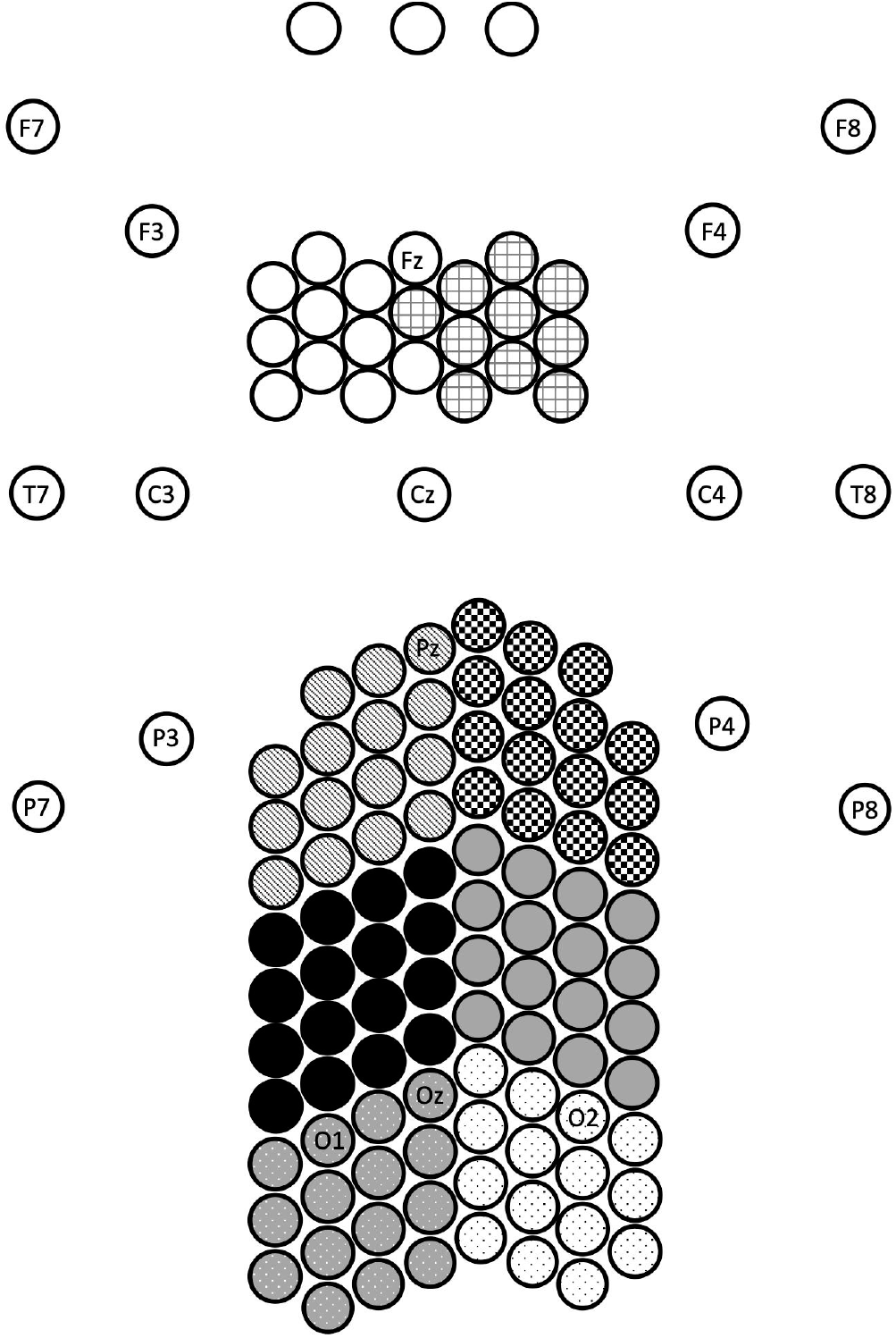
A 2D map of electrode locations. 10-20 electrode labels included for reference. Monochrome patterns used to indicate clusters used for analysis: left and right frontal, parietal, occipito-parietal, and occipital areas. Scale adjusted for illustration (Haigh et al., 2019).

To take advantage of the high-density EEG montage, for the analysis, the EEG electrodes were grouped into eight spatial clusters. These are shown in Fig. 2 as right and left occipital (dotted pattern), occipito-parietal (solid gray and black), parietal (checkerboard and stripes), and frontal electrodes (grid pattern and white) (for successful use of these clusters, see Haigh et al., 2019).

### Response time

The reaction times (RT) to the color at fixation were analyzed first. Trials with RTs longer than 1 second were excluded (∼8% of trials) as were trials with RTs that preceded the color change (∼7.5% of the trials). The remaining RTs were averaged for each participant x each modality (visual and auditory) x each stimulation frequency (4 and 6Hz).

### EEG pre-processing

EEG data were preprocessed using the EEGLAB (Delorme and Makeig, 2004) and ERPLAB (Lopez-Calderon and Luck 2014) toolboxes in MATLAB (MathWorks): (i) EEG data were re-referenced offline to the average of the two mastoid electrodes and a zero-phase Butterworth filter was used to filter the signals between 0.1-100Hz; (ii) Noisy channels were detected visually and interpolated (this was done in 0.95% of electrodes from the migraine group and 0.3% from the control group); (iii) Independent component analysis (ICA) was used to identify and remove eye-related artifacts (blinks and horizontal eye movements), and heartbeat; (iv) For the visual and auditory trials, the first 2 seconds after the stimulus onset were extracted, and, for the resting state trials, 2-second non-overlapping time intervals were extracted; and (v) These 2-second time intervals were passed through zero-phase Kaiser bandpass filters to extract 5 frequency bands of delta (0.5-3Hz), theta (4-7Hz), alpha (8-12Hz), beta (12-30Hz), and gamma (30-100Hz).

### Coherence

We estimated the Pearson’s correlation coefficients (PCCs) at zero-lag (n=0) as a measure of coherence between electrical activities of each pair of electrodes (Xand Y) for each frequency band (*f*). PCCs take values in the range of [-1,1]:

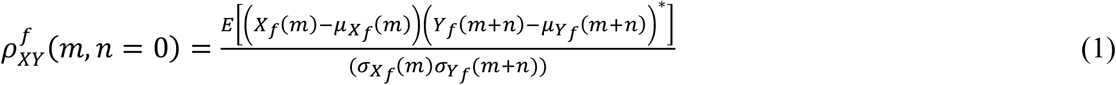

Where *μ* and *σ* are the mean and standard deviation of *X* and *Y*, which are estimated using N repeated trials, i.e., N bandpass filtered 2-second time intervals were used as sample functions of these two stochastic processes X and Y:

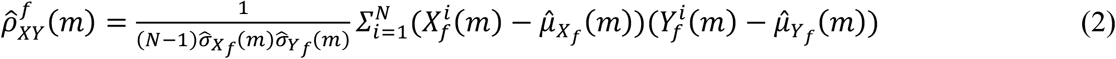

Where we have used unbiased estimators of variances, covariances, and mean of these random processes:

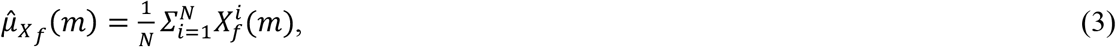

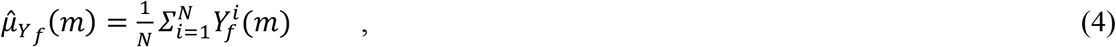

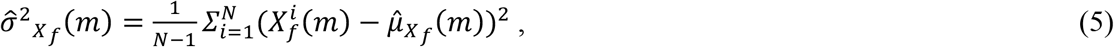

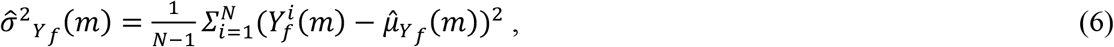

The estimated PCCs in (1) are functions of time (*m*) and frequency (*f*). We side-stepped the unrealistic stationarity assumption for EEG signals. We consider the absolute value of PCCs in our analyses, since both correlation (positive sign) and anti-correlation (negative sign) capture the coherence of activities for each pair of EEG electrodes.

#### Spatial analysis

We averaged the absolute value of estimated PCCs over 2-second time intervals to obtain a spatial map of coherence. PCCs were grouped based on the length of their corresponding links in the 2D map of electrodes (see Fig. 2), i.e., link length 1 (21-40), link length 2 (41-60), link length 3 (61-80), and link length 4 (>81, squared Euclidean distance with electrode-space unit). We excluded the links with the shortest length (<20) from the analysis to remove the spurious high correlations in nearby electrodes (Lachaux, et al., 1999). For each cluster of electrodes (see Fig. 2), we defined two measures: (i) inter-hemispheric, defined as the PCCs calculated between electrodes of a cluster and the electrodes placed on the other hemisphere, averaged for each link length, and (ii) intra-hemispheric, defined as the PCCs calculated between electrodes of a cluster and the electrodes placed on the same hemisphere, averaged for each link length.

#### Coherence normalization

Absolute value of PCCs have a different range of values for different inter-electrode distances (very small values for large links and vice versa). To ensure that this does not affect the results of the coherence analysis, we repeated the spatial coherence analysis using the normalized PCCs. Since the correlations drop exponentially as the inter-electrode distance increases (see Supplementary Fig. 9), we used an exponential regression to fit an exponential function (*ae*^*bx*^) to the absolute value of PCCs which are averaged for each inter-electrode distance. Then PCCs are normalized by this exponential function of inter-electrode distance. The results of the normalized coherence analysis are included in the Supplementary materials.

### Statistical analysis

To ascertain the effect of modality and/or stimulation frequency on the detection reaction time in migraine patients, a 3-way mixed-model ANOVA was used with 2 levels of modality (auditory and visual) and stimulation frequency (4 and 6Hz) as within-subject factors and group (migraineur and control subject) as the between-subject variable, and averaged RT as the dependent variable.

For measures of EEG coherence, we used a 7-way mixed-model ANOVA with frequency bands (delta, theta, alpha, beta, and gamma), link lengths (21-40, 41-60, 61-80, and >81), two levels of modalities (visual, and auditory), two levels of stimulation frequencies (4Hz and 6Hz), eight spatial clusters (left and right occipital, occipito-parietal, parietal, and frontal electrodes), and hemisphere (intra/inter) as within-subject factors, and group (migraineurs and control subjects) as the between-subject factor. Also, a 6-way mixed-model ANOVA was conducted on the rest dataset with five within-subject factors (EEG frequency bands, link lengths, spatial clusters, hemisphere), and group as a between-subjects factor.

Bonferroni post-hoc test with family-wise correction was used to explore any significant interactions between group and any other factor/s in the mixed-model ANOVAs. MATLAB (MathWorks) and Unixstat (Perlman, 1984) were used for all analyses in this study.

### Data availability

The anonymized raw EEG dataset of the participants in this research are made available online on KiltHub, Carnegie Mellon University’s online data repository (DOI: 10.1184/R1/12636731).

## Results

We first assessed the RTs in the visual and auditory tasks and then explored the effect of visual and auditory stimuli on the spatio-temporal pattern of hemispheric coherence. This was followed by a similar analysis of the resting-state EEG data.

### Response time

Each participant’s RTs were averaged over trials separately for stimulation frequency (4Hz and 6Hz) and modality (visual and auditory), and these factors were subjected to an ANOVA with group (migraineur and control) as the between-subject variable. Migraineurs had significantly faster response times (mean=465ms) compared to the headache-free control subjects (mean=527ms; main effect of group: F(1,35)=10.35, p<.002). There were no significant interactions of group with any of the other factors.

### Coherence

#### (i) Measures of coherence during visual and auditory stimulation

The mixed-model ANOVA revealed a five-way interaction of group x link lengths x frequency bands x modalities x stimulation frequencies (F(12,312)=1.83, p<.05), a five-way interaction of group x frequency bands x spatial clusters x stimulation frequencies x link lengths (F(84,2184)=1.71, p<.001), and a five-way interaction of group x frequency bands x modalities x hemisphere x link lengths (F(12,312)=2.72, p<.005), each of which we break down below. Additionally, there was a four-way interaction of group x spatial clusters x stimulation frequencies x hemisphere (F(7,182)=2.13, p<.05) and a four-way interaction of group x spatial clusters x modalities x link lengths (F(21,546)=2.26, p<.001), along with several lower level interactions which are all subsets of the aforementioned higher-level interactions.

To uncover the nature of the interactions, multiple post-hoc comparisons were run using a conservative Bonferroni family-wise correction. Fig. 3 shows the spatial coherence values for different modalities (auditory and visual), and different stimulation frequencies (4 and 6Hz) as a function of link length for each of the five frequency bands and groups (the five-way interaction of group x link length x frequency bands x modalities x stimulation frequencies). Based on the results, compared with controls, migraineurs showed significantly lower spatial coherence in the alpha frequency band for auditory 4Hz stimulation frequencies in large link lengths of 3 and 4 (p<.001; M=.430±.009^1^, C=.491±.009), visual 4Hz stimulation frequencies in large link length of 4 (p<1e-4; M=.351±.012, C=.423±.012), and visual 6Hz stimulation frequencies in large link length of 3 and 4 (p<.001; M=.439±.010, C=.511±.009). To summarize, migraineurs showed significantly lower long-range spatial coherence of alpha-band neural activities at 4Hz stimulation frequencies in both modalities of audition and vision, and at 6Hz stimulation frequencies in vision (but not at 6Hz audition although the trend is in the same direction).

**Figure 3.**
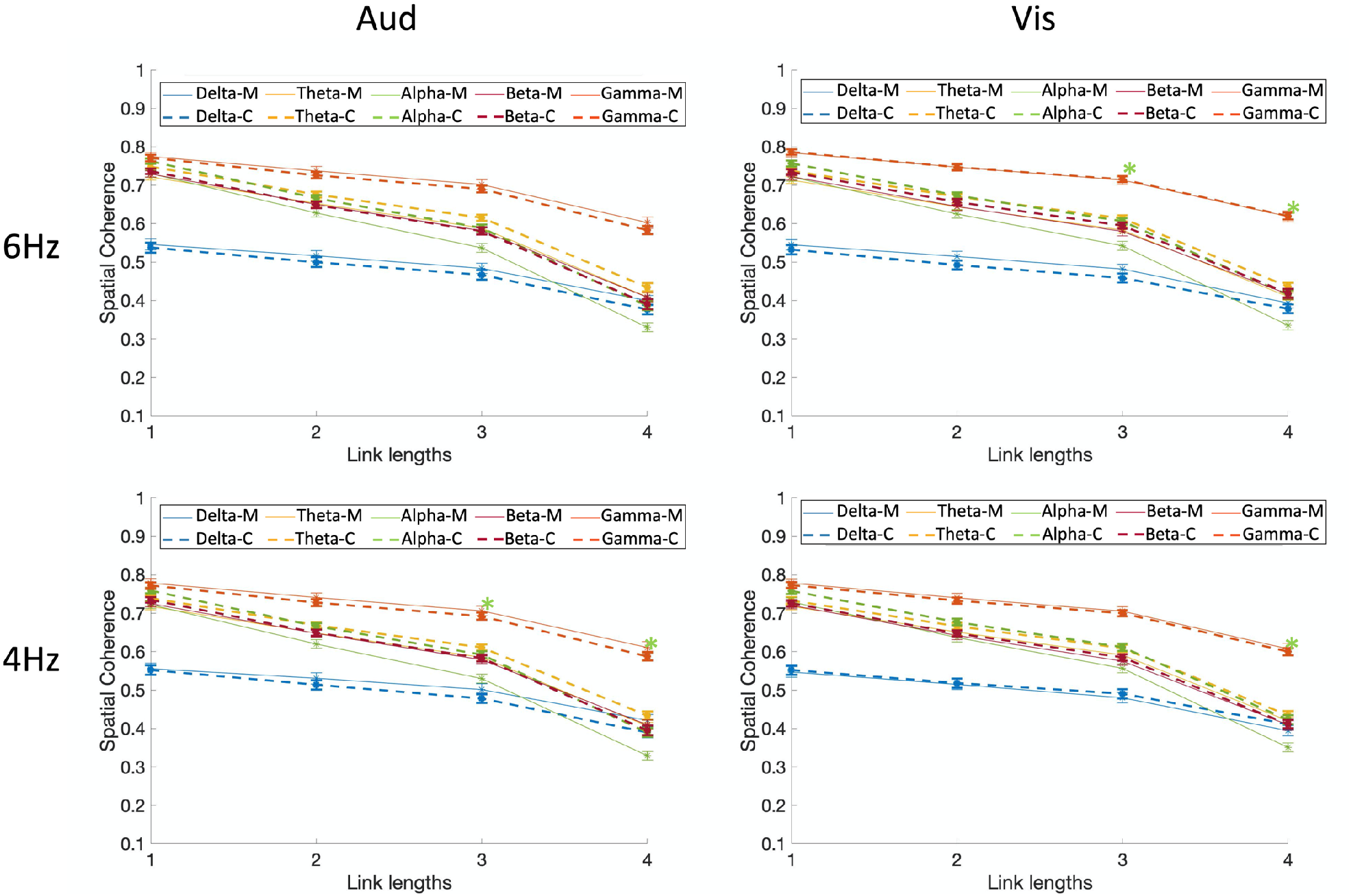
Five-way coherence interaction of group x link length x frequency bands x modalities x stimulation frequencies. Comparison of spatial coherence between migraine and controls for visual and auditory modalities, and different stimulation frequencies (4 and 6Hz) as a function of link length for each of the five frequency bands. Spatial coherence is shown using dashed lines for controls and solid lines for migraineurs. Asterisks show the significant group differences for each link length (on the x-axis) and each frequency band (colors of asterisks are matched with the frequency bands), based on Bonferroni post-hoc test (M= migraineurs, C=controls).

Fig. 4A shows the spatial coherence values for stimulation frequency of 4Hz as a function of link lengths for each of the five frequency bands, each of the eight spatial clusters, and groups (the five-way interaction of group x frequency bands x spatial clusters x stimulation frequencies x link length). Note that the data are collapsed across modality as this factor is not included in this interaction. Fig. 4B shows the interactions for stimulation frequency of 6Hz. Compared with controls, migraineurs showed significantly lower spatial coherence in the frontal clusters for the alpha frequency band with link length 3 for both stimulation frequencies (p<1e-4; M=.408±.012, C=.547±.009), and link length 2 but just for 4Hz stimulation frequency (p<1e-4; M=.484±.017, C=.620±.011), although the trend for the same finding is present in 6Hz too. These results indicate that migraineurs showed significantly lower long-range spatial coherence of alpha-band neural activities in the frontal clusters during both stimulation types (visual/auditory) and frequencies (4 and 6Hz).

**Figure 4.**
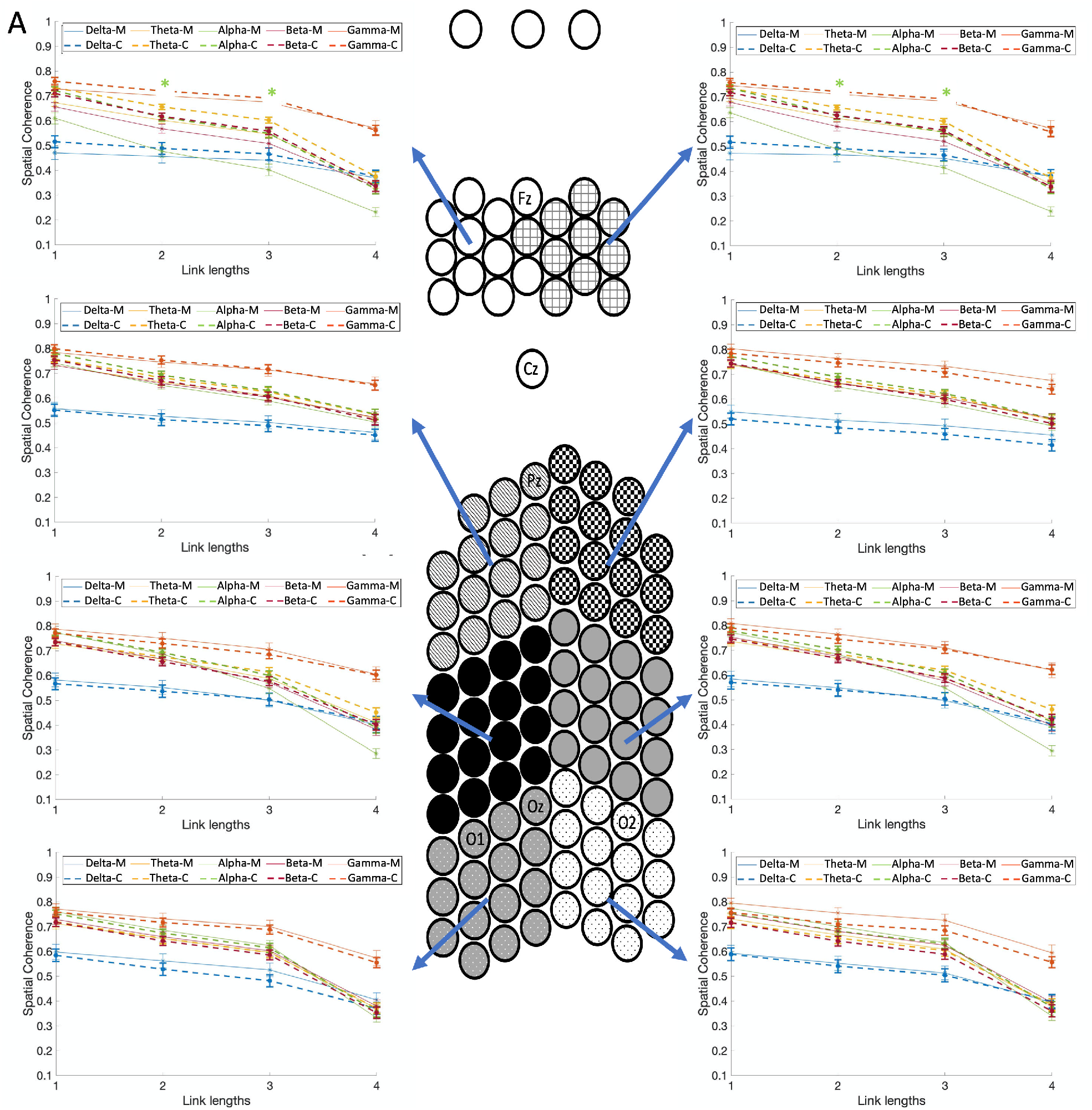
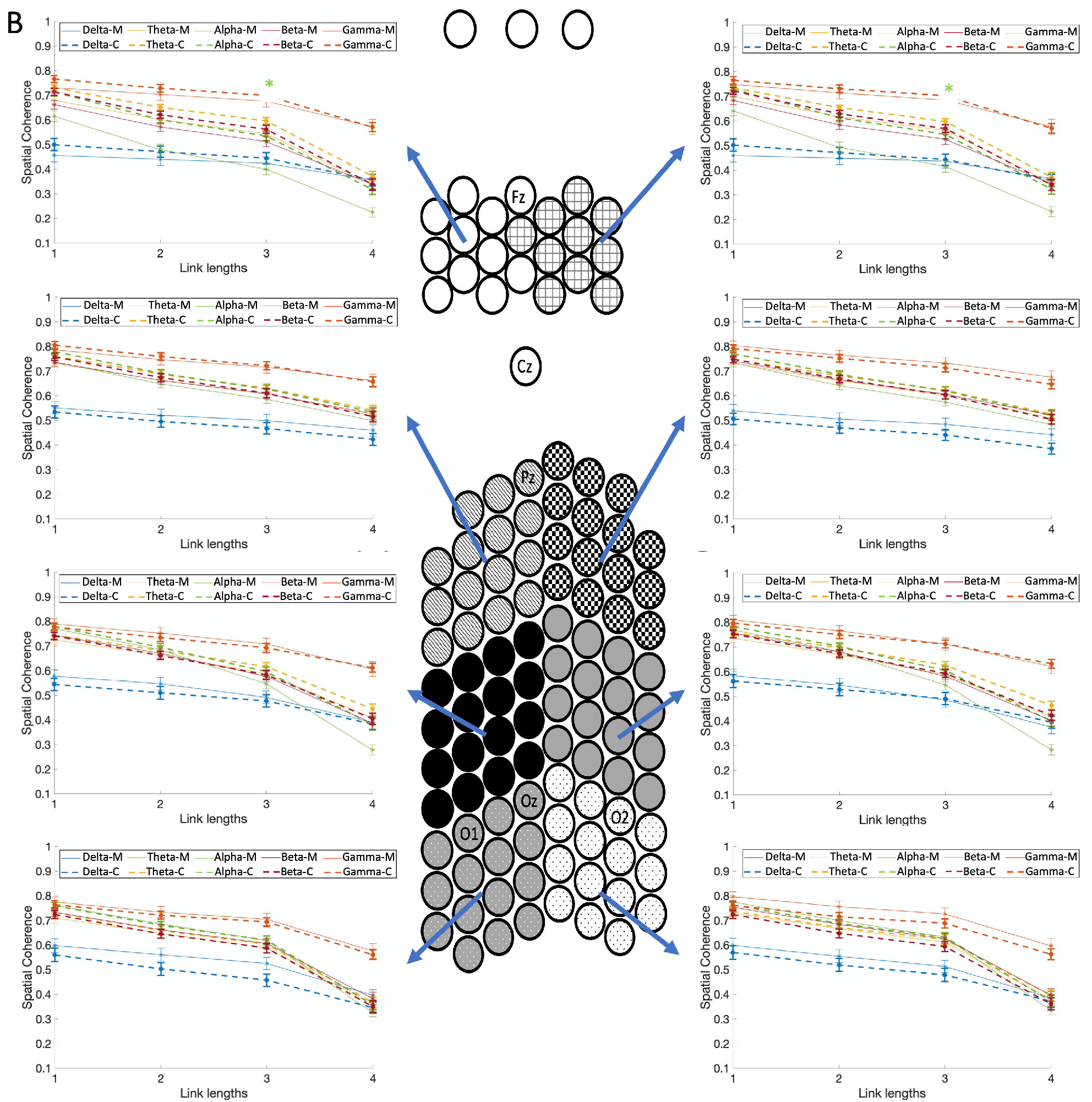
Five-way coherence interaction of group x frequency bands x spatial clusters x stimulation frequencies x link length. Comparison of spatial coherence between individuals with migraine and controls for (**A**) the stimulation frequency of 4Hz, and (**B**) 6Hz, as a function of link lengths for each of the five frequency bands, each of the eight spatial clusters, and groups (the five-way interaction of group x frequency bands x spatial clusters x stimulation frequencies x link lengths). Spatial coherence is shown using dashed lines for controls and solid lines for the migraine patients. Asterisks show the significant group differences for each link length (on the x-axis) and each frequency band (colors of asterisks are matched with the frequency bands), based on Bonferroni post-hoc test (M= migraineurs, C=controls).

Fig. 5 shows the spatial coherence values for both modalities (auditory and visual), and hemisphere (intra/inter) as a function of link length for each of the five frequency bands and groups (the five-way interaction of group x frequency bands x modalities x hemisphere x link lengths). Compared with controls, migraineurs showed significantly lower inter-hemispheric coherence in the alpha frequency band with link length 4, for both modalities (p<0.001; M=.357±.008, C=.421±.008). The migraineurs also showed significantly lower intra-hemispheric coherence in the alpha frequency bands with link length 3, for both modalities (p<1e-4; M=.505±.008, C=.574±.007), and with link length 4 for but just for visual stimulation (p<1e-5; M=.322±.012, C=.403±.012). To summarize, relative to controls, migraineurs showed significantly lower long-range inter- and intra-hemispheric coherence of alpha-band neural activities in all stimulation types (visual/auditory) and frequencies (4 and 6Hz).

**Figure 5.**
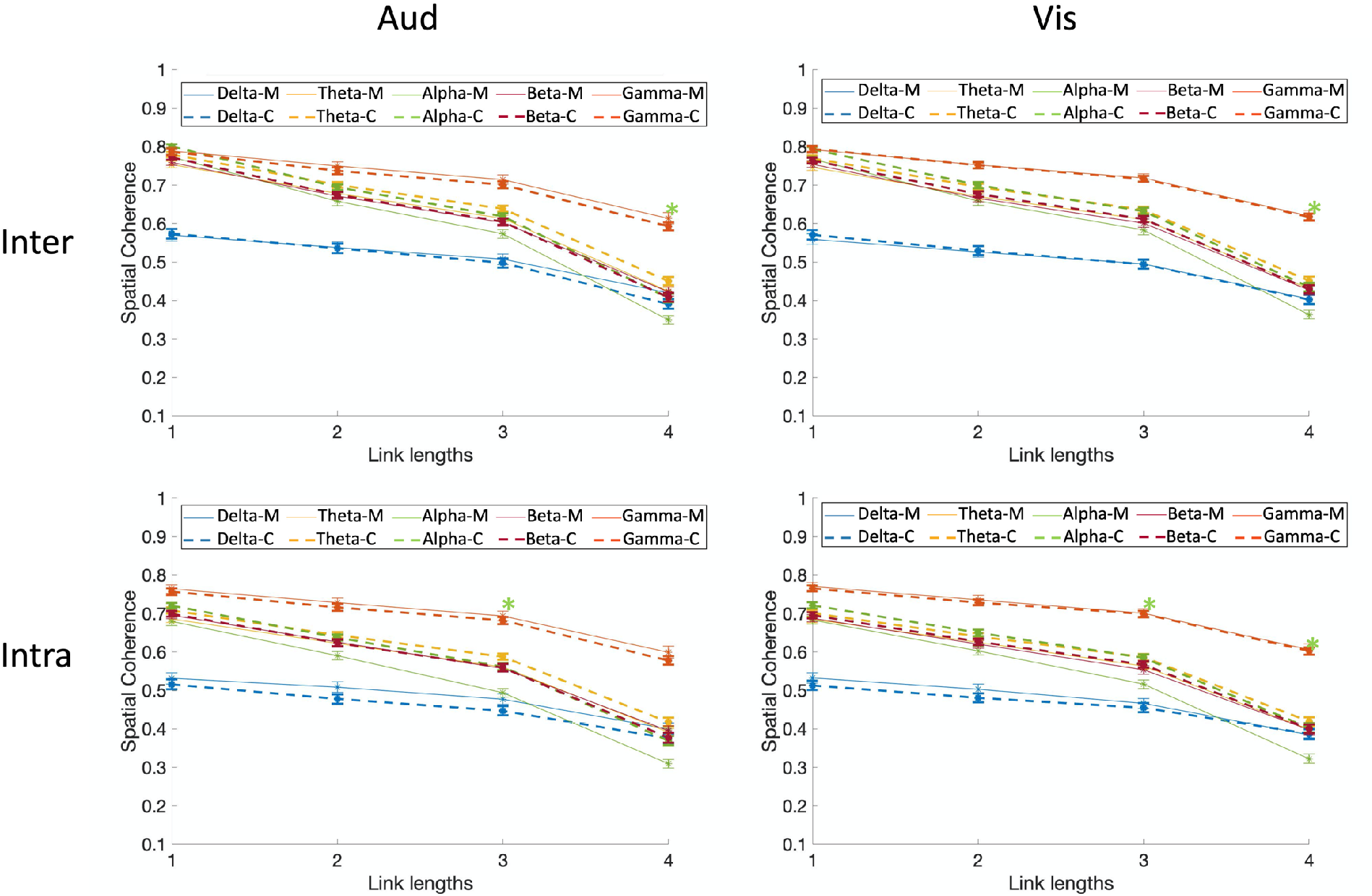
Five-way coherence interaction of group x frequency bands x modalities x hemisphere x link lengths. Comparison of spatial coherence between individuals with migraine and controls for different modalities (auditory and visual), and different hemisphere (intra/inter) as a function of link lengths for each of the five frequency bands and groups. Spatial coherence is shown using dashed lines for controls and solid lines for the migraine patients. Asterisks show the significant group differences for each link length (on the x-axis) and each frequency band (colors of asterisks are matched with the frequency bands), based on Bonferroni post-hoc test (M= migraineurs, C=controls).

Fig. 6 shows the spatial coherence values for the two stimulation frequencies (4Hz/ 6Hz) as a function of hemisphere (intra/inter) for the eight spatial clusters, and groups (the four-way interaction of group x spatial clusters x stimulation frequencies x hemisphere). Migraineurs showed significant lower coherence in the left frontal clusters for both stimulation frequencies and inter- and intra-hemisphere connections (p<0.001; M=.517±.005, C=.567±.004), and significant lower inter-hemispheric coherence in the right frontal cluster for 4Hz stimulation frequency (p<0.001; M=.539±.009, C=.586±.008). In summary, relative to controls, migraineurs showed significantly lower coherence (inter- and intra-hemispheric) in the left frontal cluster for both stimulation frequencies, and lower inter-hemispheric coherence in the right frontal cluster for 4Hz stimulation frequency, regardless of the stimulation type.

**Figure 6.**
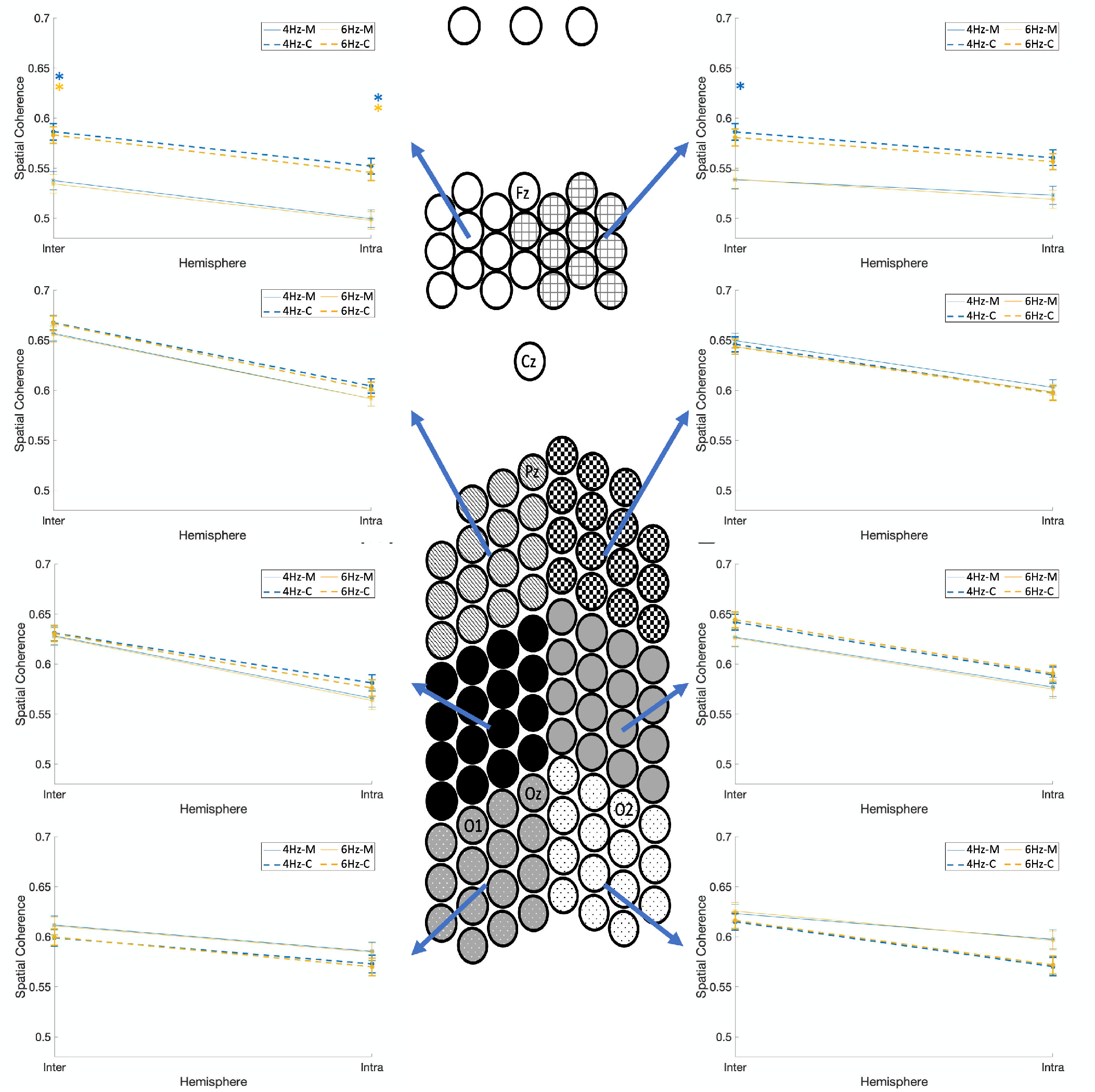
Four-way coherence interaction of group x spatial clusters x stimulation frequencies x hemisphere. Comparison of spatial coherence between individuals with migraine and controls for the two stimulation frequencies (4Hz and 6Hz) as a function of hemisphere (intra/inter) for each of the eight spatial clusters, and groups. Spatial coherence is shown using dashed lines for controls and solid lines for the migraine patients. Asterisks show the significant group differences for each hemisphere (intra/inter on the x-axis) and each stimulation frequency (colors of asterisks are matched with the stimulation frequency), based on Bonferroni post-hoc test (M= migraineurs, C=controls).

Fig. 7 shows the spatial coherence values for the two different modalities as a function of link lengths for each of the eight spatial clusters, and groups (the four-way interaction of group x spatial clusters x modalities x link lengths). Relative to controls, migraineurs showed significantly lower spatial coherence in the frontal clusters for visual stimulation and link lengths of 2 and 3 (p<0.001; M=.537±.006, C=.603±.004), and left frontal cluster for visual stimulation and smallest link length of 1 (p<0.001; M=.626±.011, C=.689±.009). In other words, migraineurs showed significant lower mid-range (link lengths 2 and 3) coherence in the frontal clusters, and lower short-range (link length 1) coherence in the left frontal cluster during visual stimulation, regardless of the stimulation frequency.

**Figure 7.**
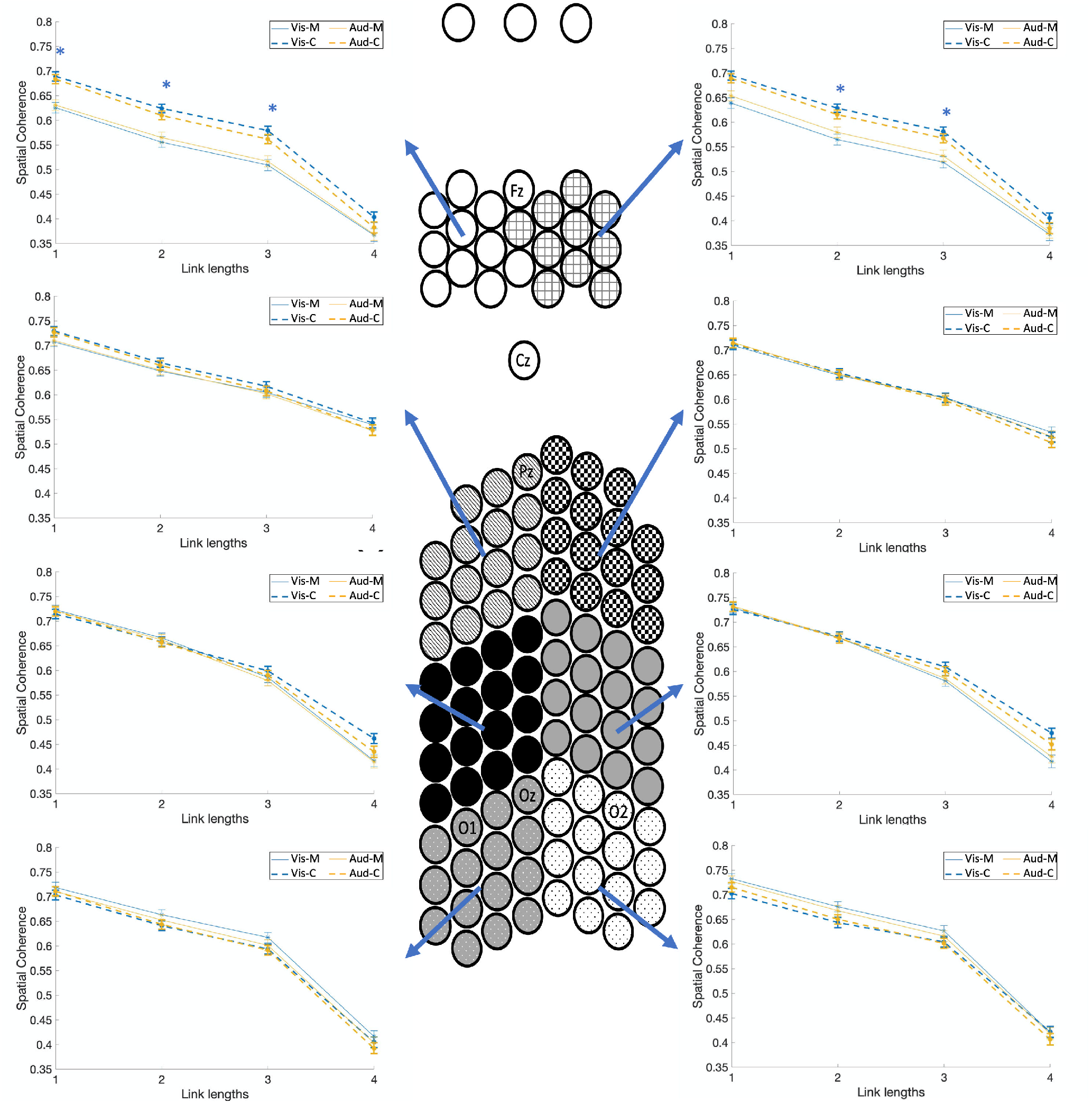
Four-way coherence interaction of group x spatial clusters x modalities x link lengths. Comparison of spatial coherence between individuals with migraine and controls for different modalities (auditory and visual) as a function of link lengths for each of the eight spatial clusters, and groups (the four-way interaction of group x spatial clusters x modalities x link lengths). Spatial coherence are shown using dashed lines for controls and solid lines for the migraine patients. Asterisks show the significant group differences for each link length (on the x-axis) and each modality (colors of asterisks are matched with the modality), based on Bonferroni post-hoc test (M= migraineurs, C=controls).

Taken together, these findings reveal that during the evoked condition, in comparison with control participants, migraineurs show lower spatial coherence usually both inter- and intra-hemispheres, more evident in frontal clusters and for longer link lengths, and evident to a greater extent in the alpha frequency range.

#### (ii) Measures of coherence during resting-state recordings

To assess whether the group differences in coherence measures were a product of sensory stimulation, we conducted the same analyses on the resting-state EEG data. A mixed-model ANOVA was conducted on the rest dataset with five within-subject factors (EEG frequency bands, link lengths, spatial clusters, hemisphere), and group as a between-subjects factor. There was a four-way interaction of group x frequency bands x spatial clusters x link length (F(84,2184)=1.51, p<.002) along with several lower level interactions which are all subsets of the aforementioned higher level interactions.

Fig. 8 shows the spatial coherence values as a function of link lengths for each of the five frequency bands, each of the eight spatial clusters, and groups (the four-way interaction of group x frequency bands x spatial clusters x link length). There were no group differences between migraineurs and headache-free controls. However, using a less conservative least significant difference (LSD) test, in comparison with controls, migraineurs showed significantly lower spatial coherence in both frontal clusters for the alpha frequency band for all link lengths (p<.04; M=.520±.009, C=.576±.007), and in the right frontal cluster for the theta frequency band for the longest link length (p<.04; M=.324±.022, C=.383±.024). In addition, in both occipito-parietal clusters, migraineurs showed significant lower coherence in the alpha frequency band for the largest link length (p<.02; M=.287±.022, C=.384±.023), and significant lower coherence in the theta frequency band for the longest link length in the right occipito-parietal cluster (p<.05; M=.396±.025, C=.469±.023).

**Figure 8.**
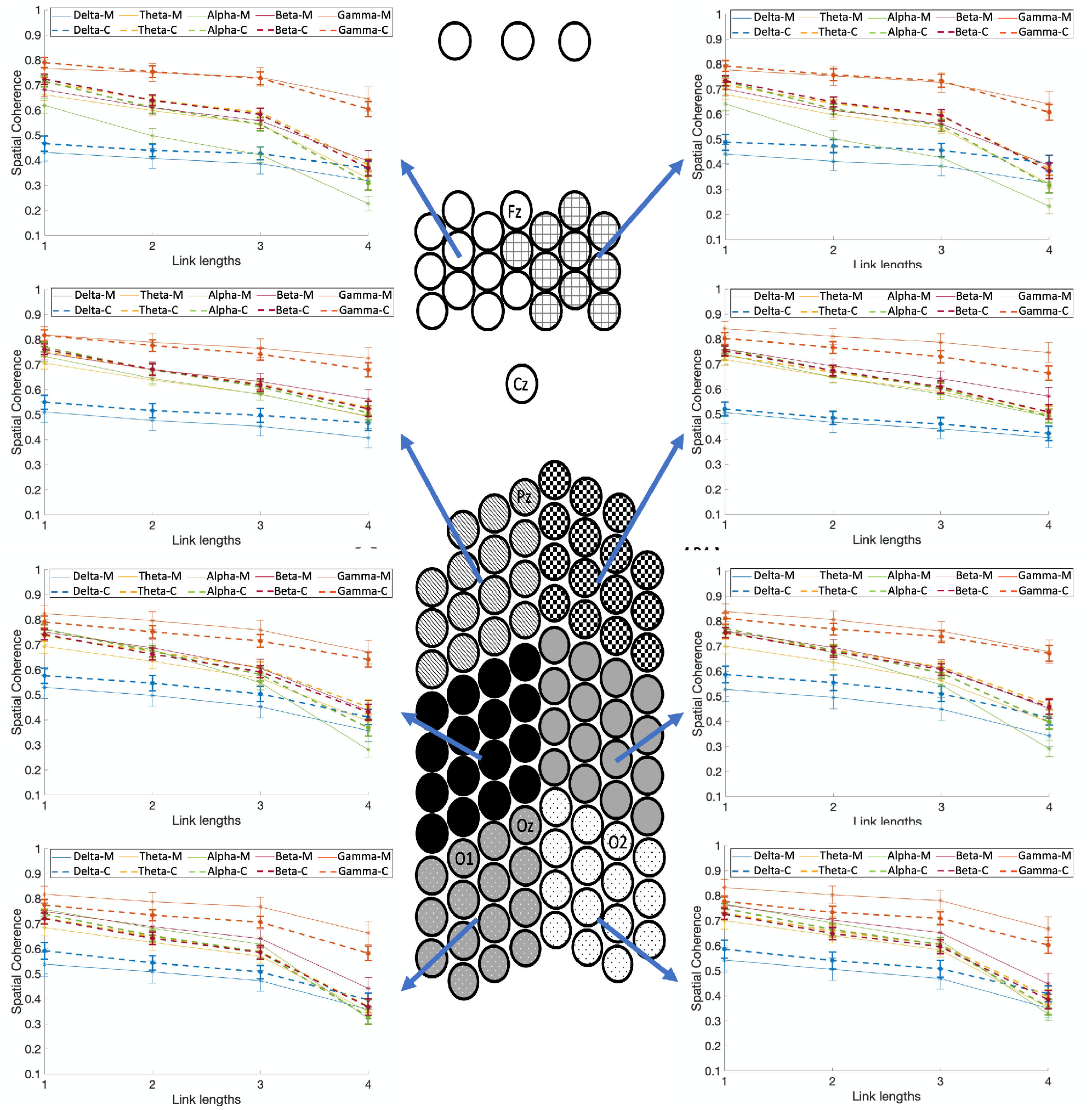
Four-way resting-state coherence interaction of group x frequency bands x spatial clusters x link lengths. Comparison of spatial coherence between individuals with migraine and controls during the resting-state recording as a function of link lengths for each of the five frequency bands, each of the eight spatial clusters, and groups (the four-way interaction of group x frequency bands x spatial clusters x link lengths). Spatial coherence are shown using dashed lines for controls and solid lines for the migraine patients. Asterisks show the significant group differences for each link length (on the x-axis) and each frequency band (colors of asterisks are matched with the frequency bands), based on Bonferroni post-hoc test (M= migraineurs, C=controls).

In summary, migraineurs did not show any significant differences in terms of spatial coherence in the resting-state in comparison with control participants. However, less conservative multiple comparisons correction revealed mainly consistent differences in coherence measures between groups in rest and in sensory-evoked scans, i.e., the frontal clusters in migraineurs showed significant lower spatial coherence in the alpha frequency band. This could be due to resting state signals (and correlations of their time courses) being weaker than signals in tasks that drive or evoke cortical responses.

The results of the normalized coherence analysis are included in the Supplementary materials, which are remarkably similar to those of the nonnormalized results reported in this section. This confirms that the different ranges of spatial coherence values for different inter-electrode distances (small values for large links and vice versa), do not affect the reported results in this study.

## Discussion

The goal of this investigation was to undertake a comprehensive evaluation of the cortical dynamics in individuals with migraine compared with headache-free controls. We examined spatial coherence (connectivity) at each of the EEG frequency bands using signals acquired from a customized ultra-high-density EEG system. Responses were measured to visual and to auditory stimulation, and at rest. Signals, both normalized and non-normalized, were compared within and between hemispheres as a function of two stimulation frequencies (4Hz and 6Hz) as well as distance between electrodes. Participants completed a color change detection at fixation in the visual and auditory stimulations trials. Several major results emerged.

Migraine participants were significantly faster at responding to the flashing cross compared to controls. This may be related to the cortex being hyper-excitable in migraine, as seen in increased visual evoked EEG to checkerboards, repetitive flashes or pattern reversal stimulation (Ambrosini et al., 2003; Aurora and Wilkinson, 2007).

Interestingly, the differences in spatial coherence networks between migraineurs and headache-free controls were evident only in the sensory-evoked scans, with no observed difference in the resting-state scans (except with a relaxed threshold). This is consistent with the heightened sensory sensitivities (photophobia and phonophobia) that are characteristic of migraine even in the interictal period (Wilkins et al., 1984; Marcus and Soso, 1989; Woodhouse et al., 1993; Huang et al., 2003; Silberstein et al., 2004; Coutts et al., 2012; Haigh et al., 2012; Joffily et al., 2016; Haigh et al., 2019). It is possible that greater sensory-evoked cortical hyper-excitability in migraine exacerbated the abnormalities in the coherence networks. Sensory-evoked signals may be more stable than signals during rest which may solve some of the discrepancies in resting-state functional connectivity in migraine (Skorobogatykh et al., 2019).

Furthermore, the spatial coherence networks were similar across visual and auditory-evoked scans, suggesting altered cortical dynamics in visual and auditory processing in migraine (De Tommaso et al., 2014; Ambrosini et al., 2017). While these data cannot directly speak to the cortical hyper-excitability in visual and auditory modalities, they do suggest that similar mechanisms that are perturbed in the visual system may also be present in the auditory system in migraine. Further exploration into the similarities and differences in auditory compared to visual functioning in migraine may help to understand the cortical changes that occur in migraine.

Surprisingly, there has been rather minimal investigation on auditory functioning in migraine, and findings of functional dynamics in resting and visual-evoked scans are rather inconsistent (Koeda et al., 1999; De Tommaso et al., 2013; Frid et al., 2020). To rectify this, we examined auditory and visual evoked signals across the five EEG frequency bands. The key results from the spatial coherence analysis, a measure of synchronization of the electro-cortical activities (Markovska-Simoska, et al., 2018), revealed that migraineurs showed significantly *lower* spatial coherence of alpha-band neural activities during the auditory and visual stimuli in comparison with the controls. This profile was especially evident in lower to mid-range distances (not too large, not too short) between the frontal clusters of scalp electrodes and other clusters especially during the visual stimulation. The migraineurs also showed significantly lower coherence between the right frontal cluster and the left hemisphere (inter-hemisphere coherence) during the 4Hz visual and auditory stimuli. Spatial coherence did not differ substantially between the groups in the resting state data when conservative family-wise correction was imposed, although there were some trends to group differences in frontal and occipital clusters at lower significance thresholds. Desynchronization of mid-range connections (lower mid-range coherence) in the alpha band, suggests greater functional activity (Singh et al., 2003; Brookes et al., 2005; Zumer et al., 2010), and is consistent with the cortex being hyper-excitable in migraine (Haigh et al., 2018).

Desynchronization in migraine may be explained by thalamocortical dysrhythmia: underactivity in thalamic nuclei that results in reduced neural synchrony across the brain (Llinas, 1988), although is often identified through desynchronization in low-frequency oscillations (theta range). Thalamocortical dysrhythmia has been linked to migraine as a potential cause for the cortical hyper-excitability and sensory disruptions (De Tommaso et al., 2014; Hodkinson et al., 2016b). However, the desynchronization in alpha band signals in migraine appears to be in contradiction with reported increased phase synchronization in alpha band in interictal migraine without aura patients in (De Tommaso et al., 2013). This could be due to the majority of our migraine participants experiencing auras (9/14), and so there may be an impact of aura that generates different neural signatures in the alpha response. Our current migraine sample is too small to assess the specific effect of aura on coherence, but this may be an interesting avenue for future study.

Identifying atypical electrophysiology in individuals with migraine is useful from both a basic science and a translational perspective. Uncovering the alterations in cortical dynamics can ascertain which neural signatures are related to different disorders. Spatial coherence metrics such as those identified here have the potential to diagnose migraineurs (Frid et al., 2020). For example, lower connectivity with the theta band was successfully used to predict group membership using a classifier. This reduction in connectivity is consistent with the findings we have obtained. Changes in spectral power may also be able to predict migraine onset and self-testing at home with a portable, relatively inexpensive EEG system is becoming feasible (Martins et al., 2020). Further exploration into how connectivity changes over the course of the migraine cycle will help with identification of when the next migraine attack will occur allowing for more targeted prophylactic treatment (Loder and Rizzoli, 2006).

In addition, identifying the cortical dynamics that are unique to migraine will help ascertain the mechanisms that contribute to the migraine pathogenesis. These mechanisms could then lead to targeted treatments to reduce the severity of, or perhaps prevent, migraine (Coppola et al., 2016a; May and Schulte, 2016). Using interventions that can disrupt migraine-related network activity and decrease cortical excitability, such as transcranial direct current stimulation (tDCS) (Wickmann et al., 2015), can offer at-home non-pharmacological prophylactic treatments. Migraine can also occur or become appreciably worse after trauma (termed post-traumatic headache), particularly after a mild traumatic brain injury or concussion (Dwyer, 2018). Identifying the mechanisms that contribute to migraine can help identify the changes in the brain post-trauma, potentially leading to improved treatment.

This study has several limitations. First, as mentioned earlier, our study includes a small number of migraine patients without aura, which makes it difficult to discern differences between mingraineurs with and without aura. For this reason, we have pooled together all the participants and did not compare these two groups separately, but further comparison of these groups is clearly warranted. Second, while having high-density of scalp electrodes in patches over occipital, parietal, and frontal areas was appropriate for assessing visual and auditory stimulation, some spatial sensitivity may have been lost for the rest scan. For further analysis of coherence equally across the scalp, an EEG cap with high density electrode coverage across the head is required. Third, we only used two stimulation frequencies in our study. Using a wider range of stimulation frequencies may reveal more information on differences of cortical coherence between migraineurs and headache-free controls. We do note, however, that the findings were not that dissimilar for the two stimulation frequencies. A final consideration is that we were unable to control the time since the last migraine attack, the time to the next attack, or the number of years over which the individual suffered from migraines. Previous studies have shown that the general time course of migraine attacks normalizes neural functioning during the attack and that the abnormalities increase with longer time since attack (Judit et al., 2000; Stankewitz et al., 2011; Coppola et al., 2013; Coppola et al., 2014; Coppola et al., 2015; Coppola et al., 2016b; Cortese et al., 2017; Deen et al., 2017; Mehnert et al., 2019). Assessing changes in spatial coherence over the migraine cycle may highlight the predictive power of these network changes preceding a migraine attack (De Tommaso et al., 2014).

In summary, migraineurs evinced significantly lower mid-range cortical coherence of alpha-band neural activities in the frontal clusters during the sensory-evoked recording (auditory and visual stimuli) compared to headache-free controls. However, no substantial cortical coherence difference was observed between groups in the resting state scans. In this study, we tried to reconcile the conflicting literature on migraine cortical connectivity through a comprehensive and complex cortical coherence analysis in interictal migraine under different types of stimuli, as well as in resting state situation. We also considered multiple factors that are hypothesized to have an effect on the cortical coherence alterations in migraine. The observed abnormalities in desynchronized neural activity across the cortex in response to sensory stimulation in this study may be explained by thalamocortical dysrhythmia that has been associated with migraine (De Tommaso et al., 2014; Hodkinson et al., 2016b). Further studies to identify the underlying mechanisms of cortical coherence and sensory processing abnormalities could lead to improved treatments for migraine patients.

## Data Availability

https://doi.org/10.1184/R1/12636731

## Acknowledgments

We thank Dr. Shawn Kelly, Dr. Jeff Weldon, and Ritesh Kumar for their assistance in constructing the EEG cap, and Praveen Venkatesh, Chaitanya Goswami, and Prof Michael Tarr for useful discussions.

## Author contributions

A.C. did the conception and design, acquisition, analysis, and interpretation of data, created new code to use in work, and wrote and revised manuscript. M.B. and S.M.H. did the conception and design, interpretation of data, and revised the manuscript. S.M.H. did the data collection. P.G. did the analysis and interpretation of data and revised the manuscript..

## Supplementary material

### Coherence normalization

Because the absolute value of spatial coherence has different ranges of values for different inter-electrode distances (small values for large links and vice versa), we repeated the spatial coherence analysis using the normalized coherence values to confirm that the reported results are not affected by this potential confound. The exponential function (*ae*^*bx*^) which was fitted to the averaged absolute value of PCCs was found to be a = 0.75, b = −3.4 × 10^−9^, with 95% confidence interval of *a* ∈ [0.73,0.77] and *b* ∈ [−3.5, −3.2] × 10^−9^. Supplementary Fig. 1 shows the averaged absolute value of PCCs and the fitted curve as functions of inter-electrode distances (squared Euclidean distance with electrode-space unit in 2D map of Fig. 2).

All absolute values of PCCs are divided by this function at each inter-electrode distance and the spatial coherence analyses were repeated. Compared to the results in part (i) of this section, the normalized coherence showed some new interactions: a new five-way interaction of group x frequency bands x stimulation frequencies x hemisphere x link lengths (F(12,312)=2.18, p<.02), a new four-way interaction of group x spatial clusters x frequency bands x hemisphere (F(28,728)=1.64, p<.03), and a new four-way interaction of group x spatial clusters x frequency bands x stimulation frequencies (F(28,728)=2.12, p<.001), along with several lower level interactions which are all subsets of the aforementioned higher level interactions. However, unlike non-normalized coherence, there was no five-way interaction of group x link lengths x frequency bands x modalities x stimulation frequencies, and no five-way interaction of group x frequency bands x spatial clusters x stimulation frequencies x link lengths. We have included the figures of these interactions in the Supplementary Fig. 2-8.

In comparison with controls, migraineurs showed significantly lower spatial coherence in the frontal clusters for the alpha frequency band, for all stimulation frequencies (p<1e-8; M=.728±.011, C=.925±.009). In addition, similar to the nonnormalized coherence results, migraineurs showed significantly lower inter- and intra-hemispheric coherence in the left frontal clusters for all stimulation frequencies (p<.001; M=.883±.008, C=.963±.006), and significant lower inter-hemispheric coherence in the right frontal cluster for 4Hz stimulation frequency (p<.001; M=.923±.016, C=1.000±.013). Migraineurs also showed significantly lower inter-hemispheric coherence in the alpha frequency band with link length 4, for all modalities and stimulation frequencies (p<1e-5; M=.733±.016, C=.874±.016), and lower intra-hemispheric coherence in the alpha frequency bands with link lengths 3 and 4, for all modalities and stimulation frequencies (p<.001; M=.741±.011, C=.872±.011). Unlike nonnormalized coherence analysis, migraineurs showed a significant lower coherence in the right occipito-parietal cluster with the largest link length 4, during the visual stimuli (p<.001; M=.842±.027, C=.960±.021). Similar to the nonnormalized coherence results, no significant group difference was observed in the rest state.

Taken together, the results of the normalized coherence analyses are remarkably similar to those of the nonnormalized results with the same preponderance of differences in frontal clusters (and to some extent in occipital, especially in visual condition), and in the alpha band frequency, and largely independent of modality and independent of stimulation frequency. The same tendency for the effects to emerge in the mid-long range link lengths and more inter-than intra-hemispheric was also mirrored in the two datasets. Following are the figures of the normalized coherence analysis:

**Figure 1.**
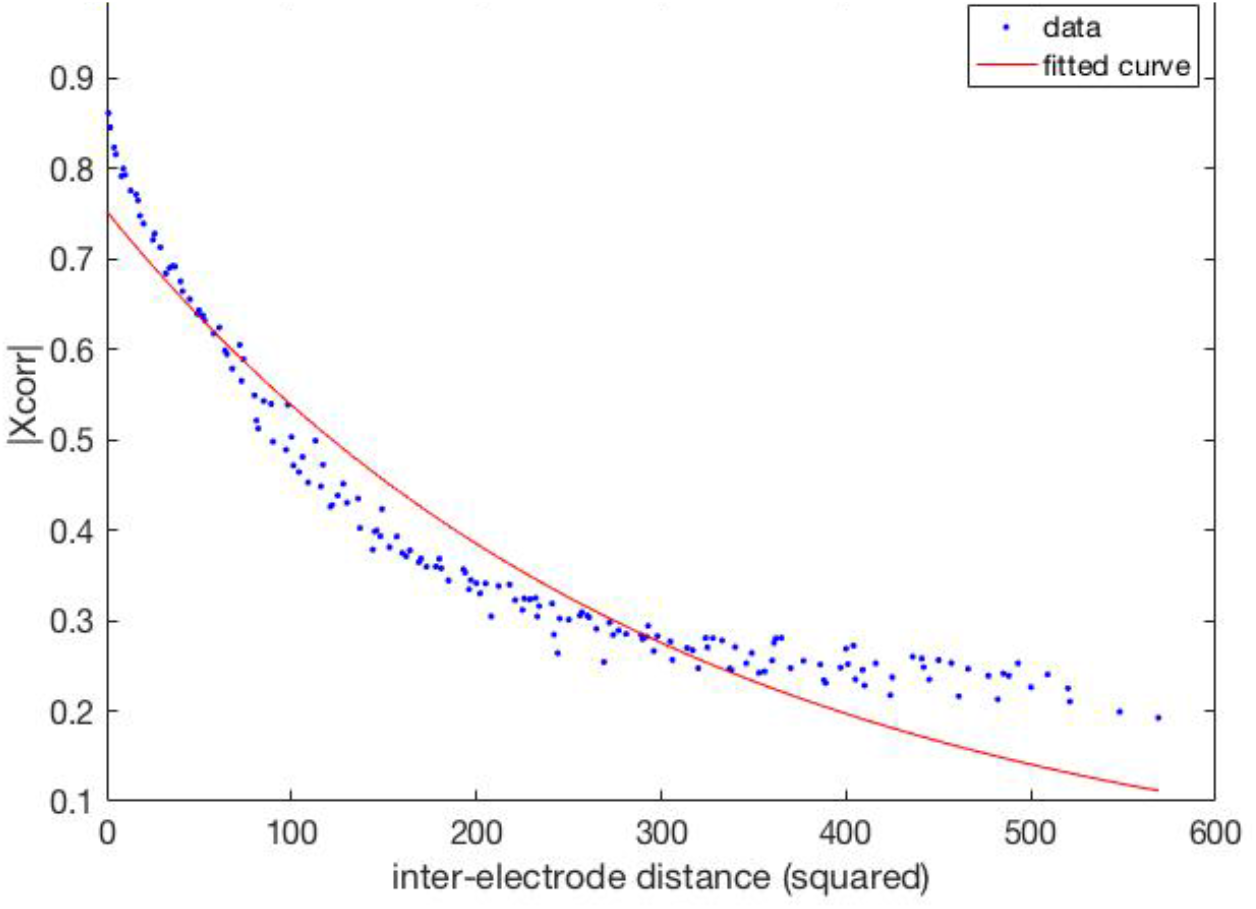
Coherence normalization. An exponential function was used (red curve) to fit to the averaged absolute value of PCCs (blue dots) as functions of inter-electrode distances (squared Euclidean distance with electrode-space unit in 2D map of electrodes shown in Fig. 2).

**Figure 2.**
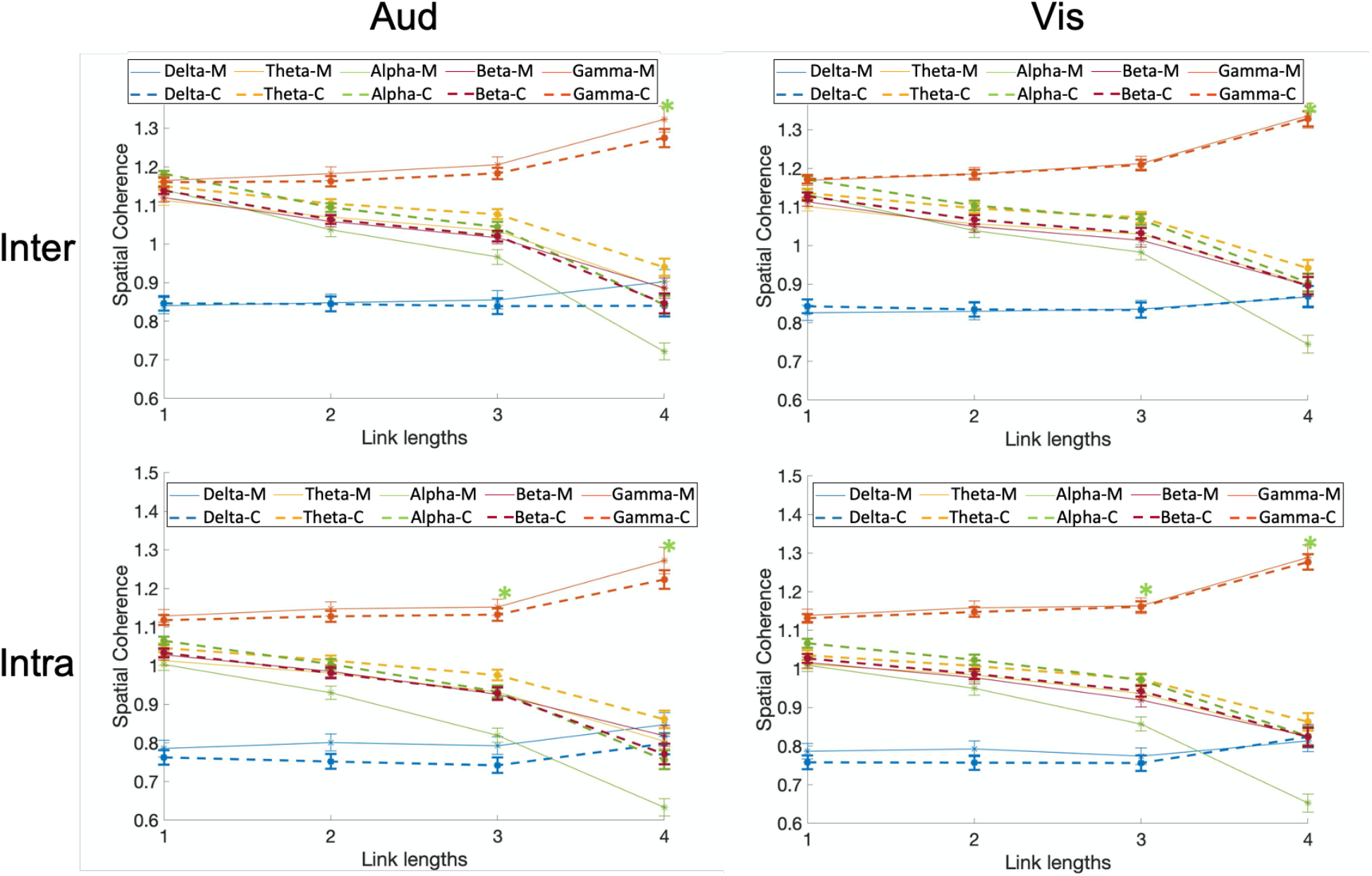
Five-way coherence interaction of group x frequency bands x modalities x hemisphere (intra/inter) x link lengths. Comparison of normalized spatial coherence between individuals with migraine and controls for different modalities (auditory and visual), and different hemisphere (intra/inter) as a function of link lengths for each of the five frequency bands and groups (the five-way interaction of group x frequency bands x modalities x hemisphere (intra/inter) x link lengths). Spatial coherence is shown using dashed lines for controls and solid lines for the migraine patients. Asterisks show the significant group differences for each link length (on the x-axis) and each frequency band (colors of asterisks are matched with the frequency bands), based on Bonferroni post-hoc test (M= migraineurs, C=controls).

**Figure 3.**
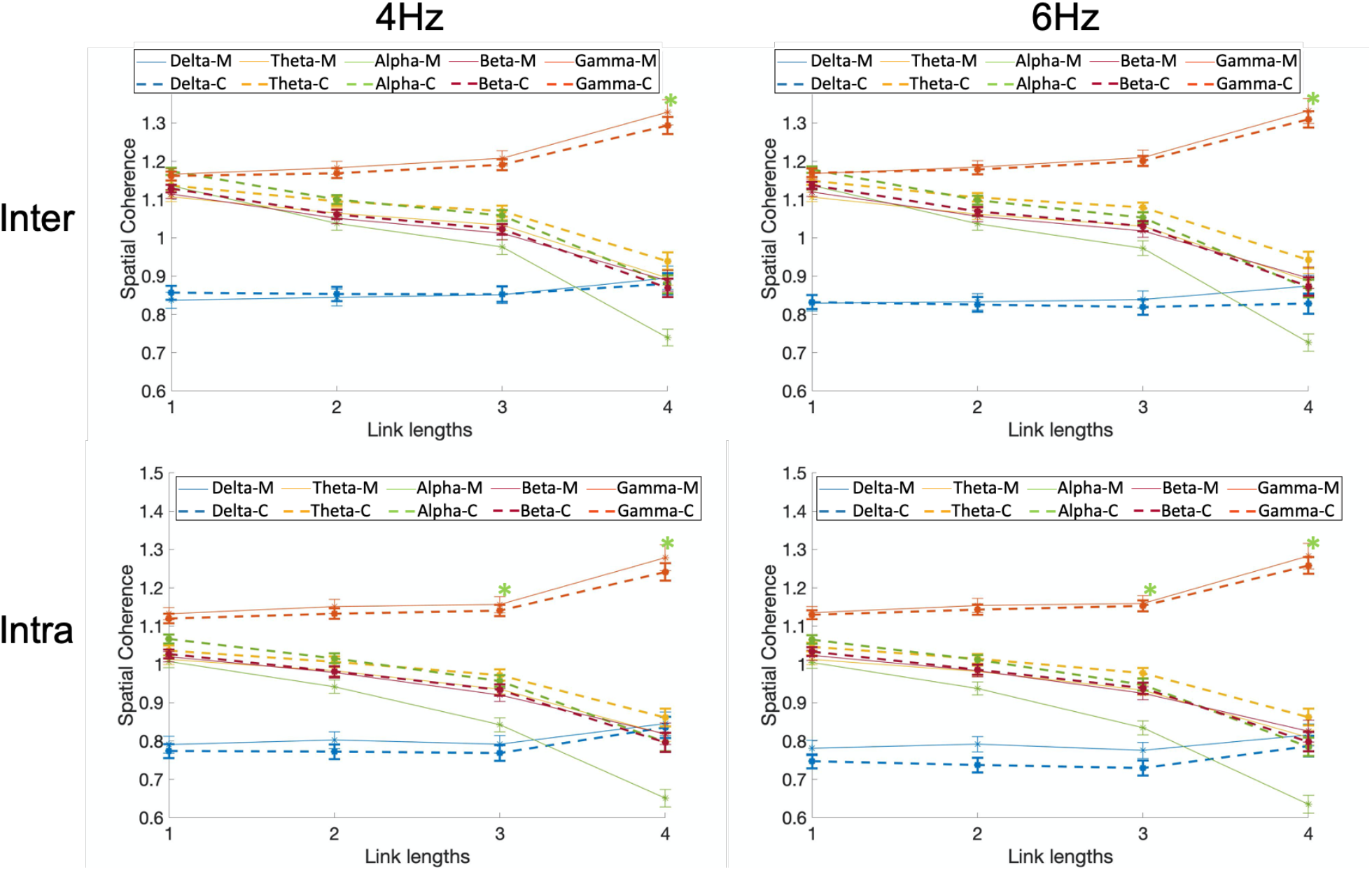
Five-way coherence interaction of group x frequency bands x stimulation frequency x hemisphere (intra/inter) x link lengths. Comparison of normalized spatial coherence between individuals with migraine and controls for different stimulation frequencies (4 and 6Hz), and different hemisphere (intra/inter) as a function of link lengths for each of the five frequency bands and groups (the five-way interaction of group x frequency bands x stimulation frequency x hemisphere (intra/inter) x link lengths). Spatial coherence is shown using dashed lines for controls and solid lines for the migraine patients. Asterisks show the significant group differences for each link length (on the x-axis) and each frequency band (colors of asterisks are matched with the frequency bands), based on Bonferroni post-hoc test (M= migraineurs, C=controls).

**Figure 4.**
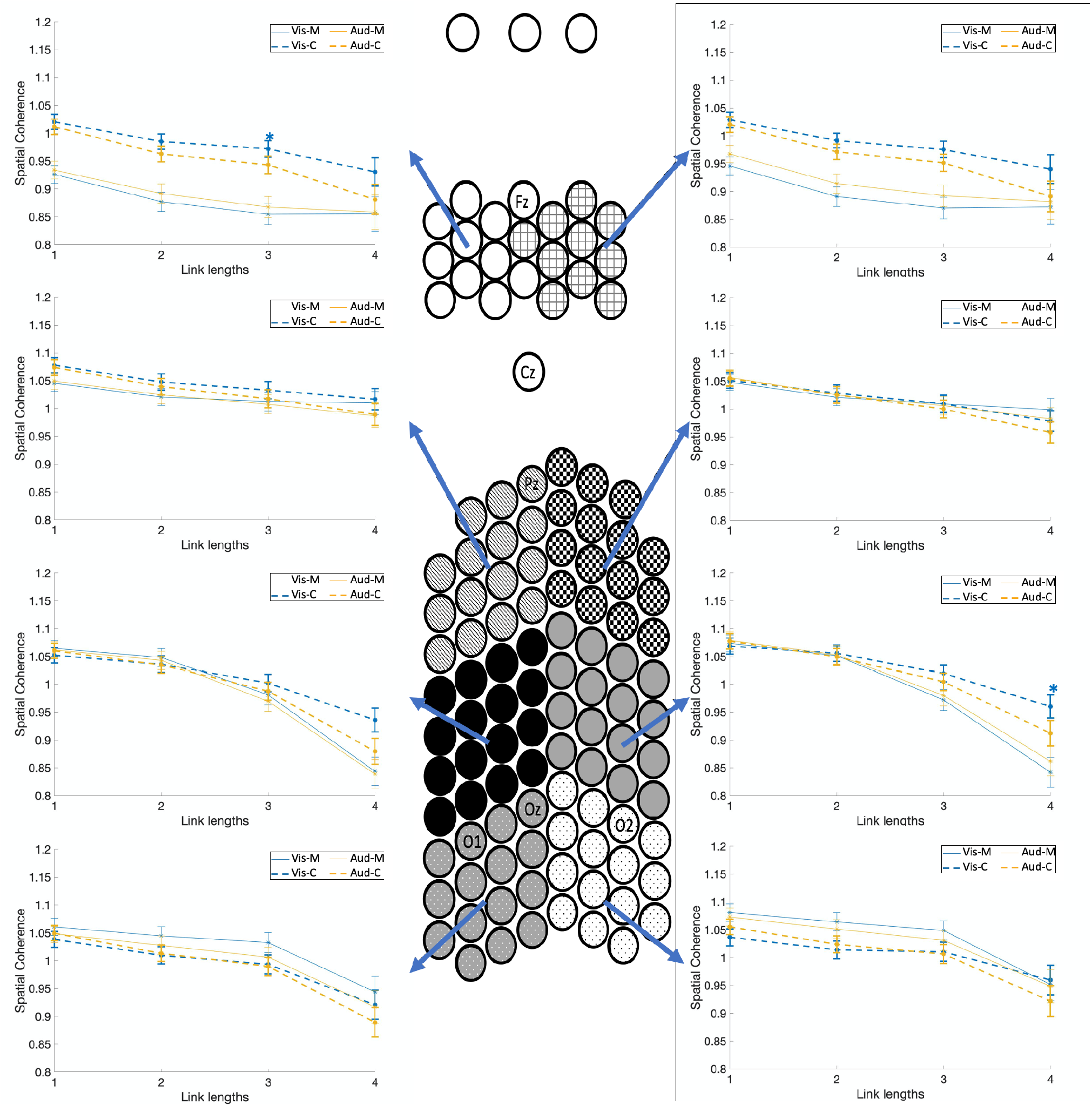
Four-way coherence interaction of group x spatial clusters x modalities x link lengths. Comparison of normalized spatial coherence between individuals with migraine and controls for different modalities (auditory and visual) as a function of link lengths for each of the eight spatial clusters, and groups (the four-way interaction of group x spatial clusters x modalities x link lengths). Spatial coherence are shown using dashed lines for controls and solid lines for the migraine patients. Asterisks show the significant group differences for each link length (on the x-axis) and each modality (colors of asterisks are matched with the modality), based on Bonferroni post-hoc test (M= migraineurs, C=controls).

**Figure 5.**
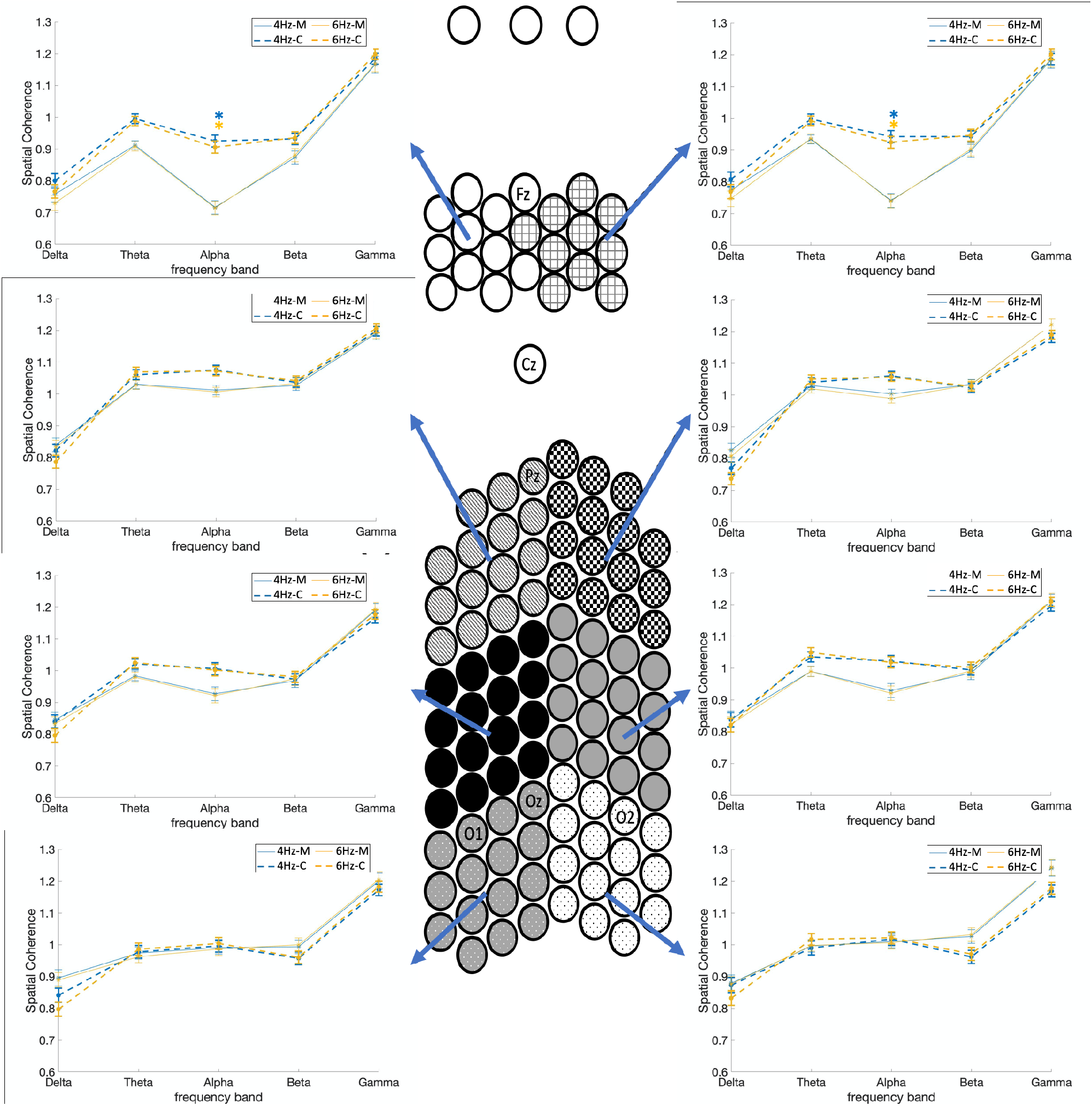
Four-way coherence interaction of group x spatial clusters x stimulation frequency x frequency bands. Comparison of normalized spatial coherence between individuals with migraine and controls for different stimulation frequencies (4 and 6Hz) as a function of frequency band for each of the eight spatial clusters, and groups (the four-way interaction of group x spatial clusters x stimulation frequency x frequency bands). Spatial coherence are shown using dashed lines for controls and solid lines for the migraine patients. Asterisks show the significant group differences for each frequency band (on the x-axis) and each stimulation frequency (colors of asterisks are matched with the stimulation frequency), based on Bonferroni post-hoc test (M= migraineurs, C=controls).

**Figure 6.**
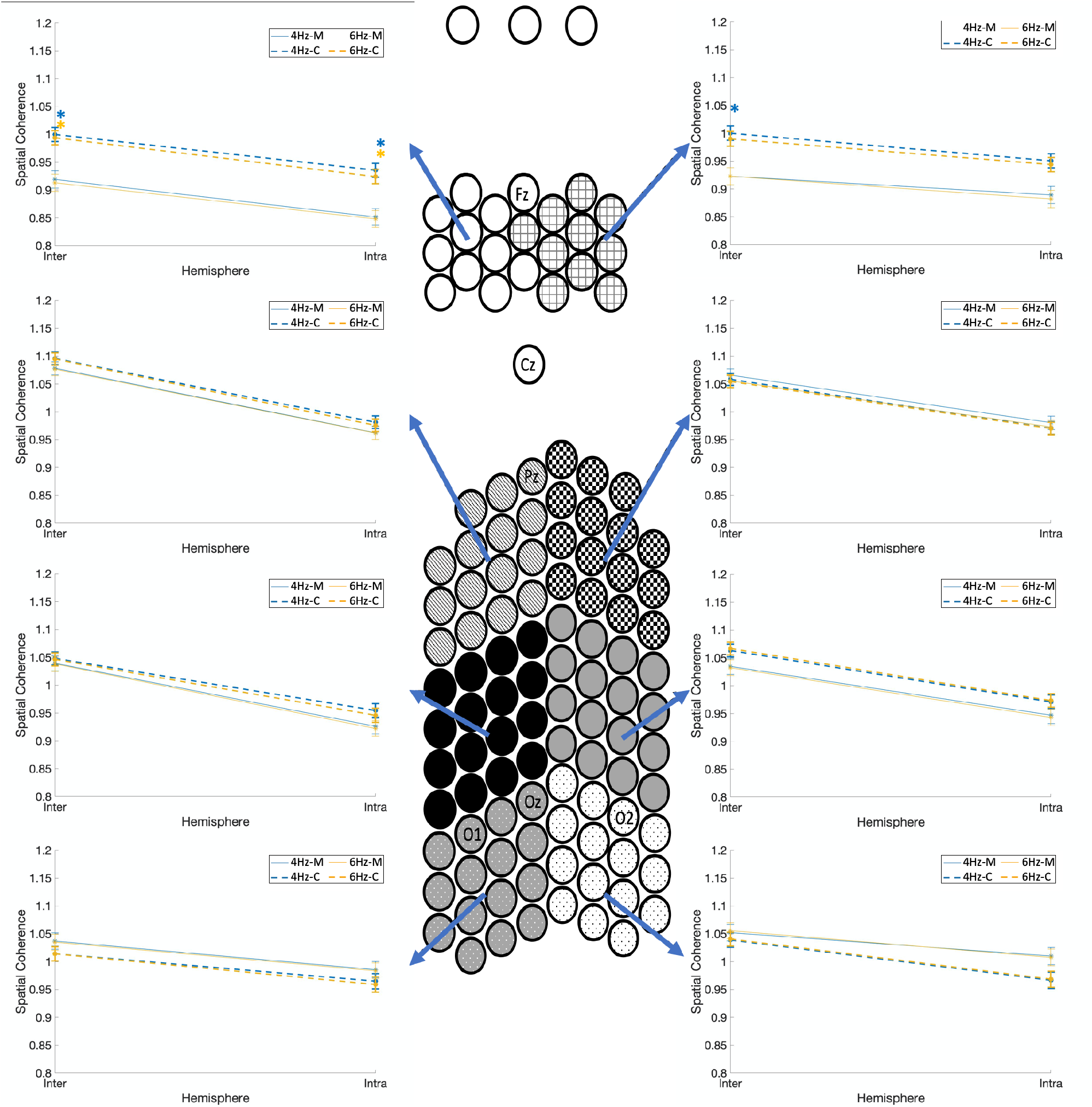
Four-way coherence interaction of group x spatial clusters x stimulation frequencies x hemisphere (intra/inter). Comparison of normalized spatial coherence between individuals with migraine and controls for the two stimulation frequencies (4Hz and 6Hz) as a function of hemisphere (intra/inter) for each of the eight spatial clusters, and groups (the four-way interaction of group x spatial clusters x stimulation frequencies x hemisphere (intra/inter)). Spatial coherence is shown using dashed lines for controls and solid lines for the migraine patients. Asterisks show the significant group differences for each hemisphere (intra/inter on the x-axis) and each stimulation frequency (colors of asterisks are matched with the stimulation frequency), based on Bonferroni post-hoc test (M= migraineurs, C=controls).

**Figure 7.**
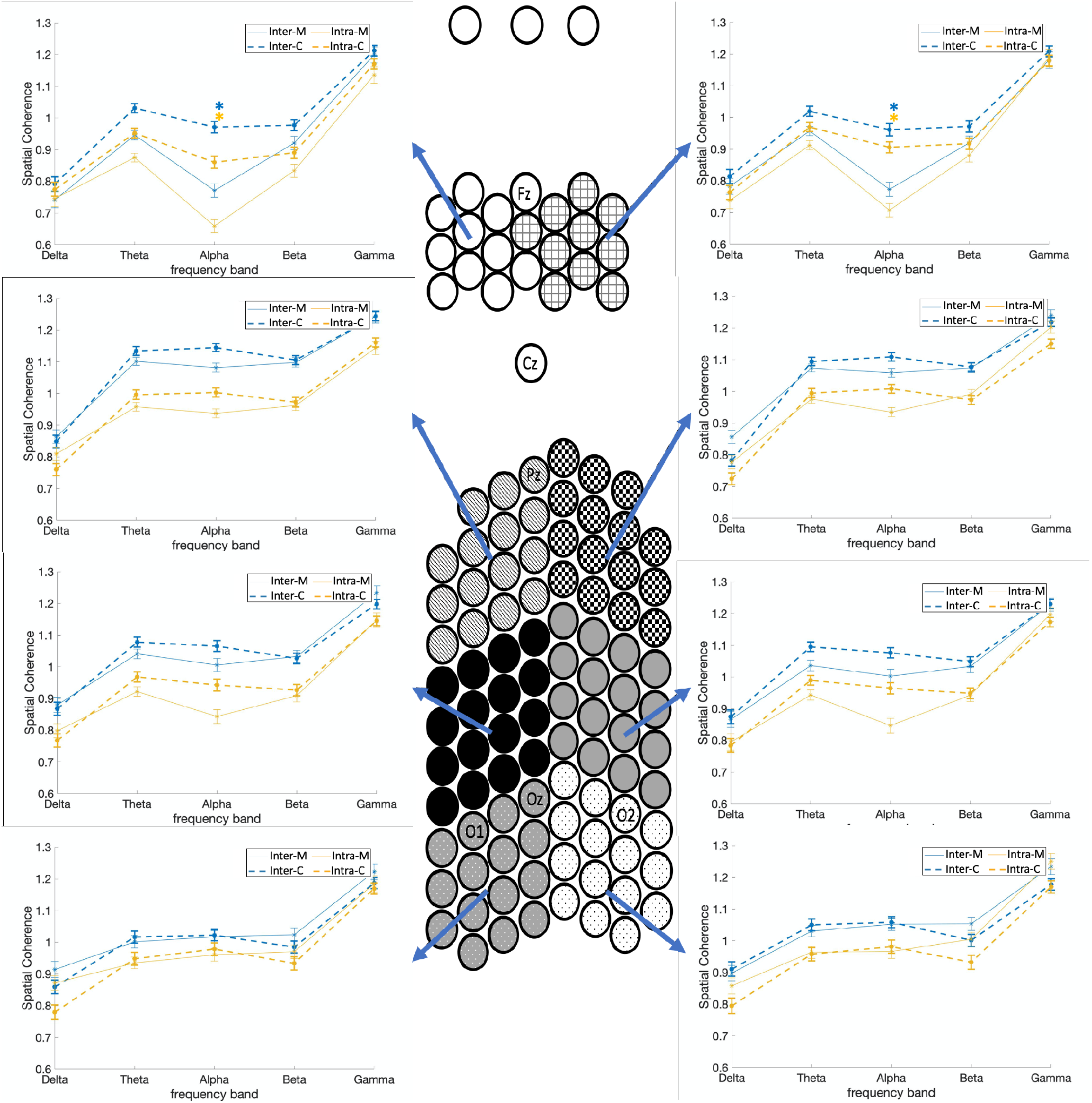
Four-way coherence interaction of group x spatial clusters x hemisphere x frequency bands). Comparison of normalized spatial coherence between individuals with migraine and controls for different hemispheres (inter/intra) as a function of frequency band for each of the eight spatial clusters, and groups (the four-way interaction of group x spatial clusters x hemisphere x frequency bands). Spatial coherence are shown using dashed lines for controls and solid lines for the migraine patients. Asterisks show the significant group differences for each frequency band (on the x-axis) and each hemisphere (colors of asterisks are matched with the hemisphere (inter/intra)), based on Bonferroni post-hoc test (M= migraineurs, C=controls).

**Figure 8.**
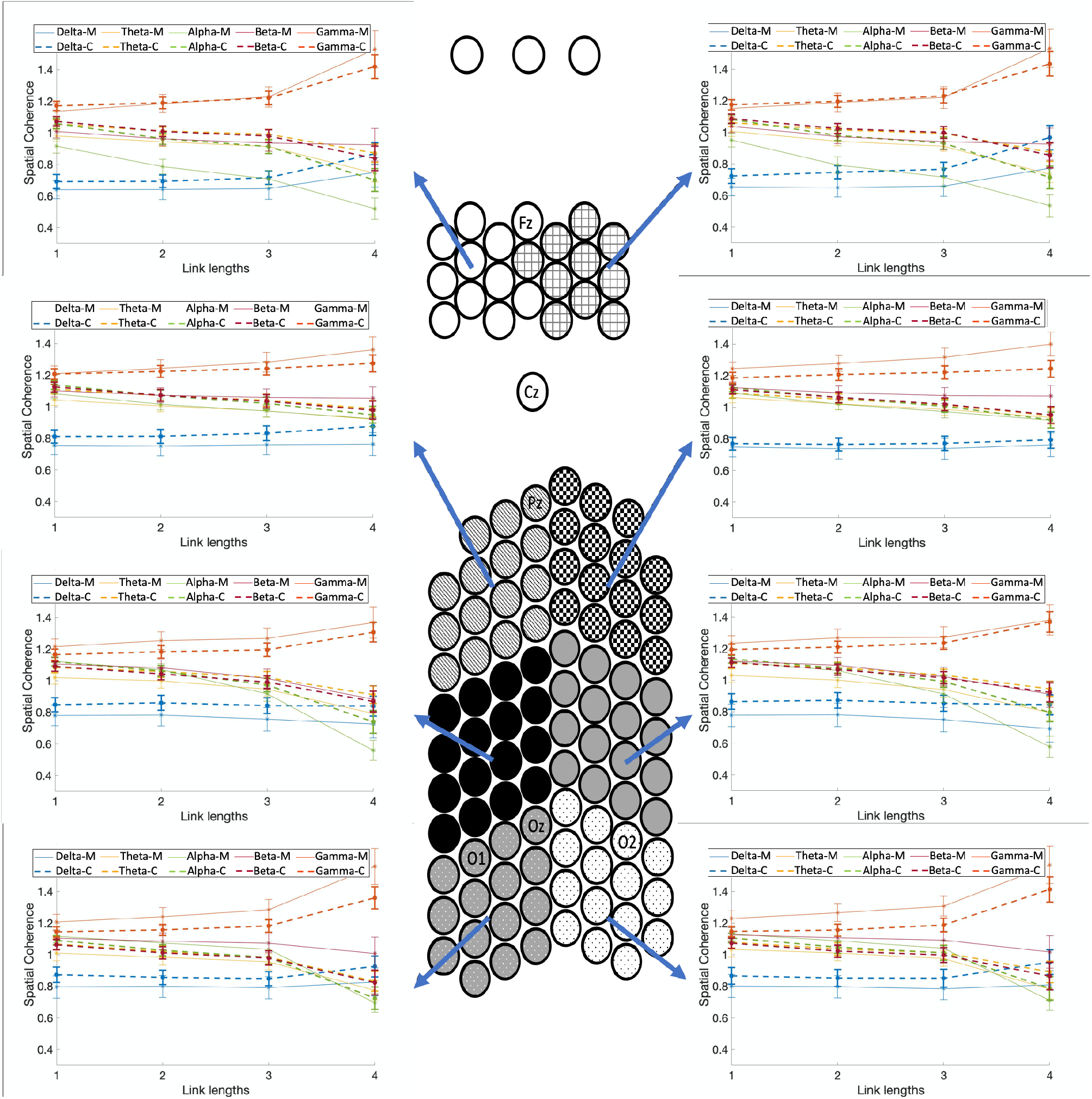
Four-way resting-state coherence interaction of group x frequency bands x spatial clusters x link lengths). Comparison of normalized spatial coherence between individuals with migraine and controls during the resting-state recording as a function of link lengths for each of the five frequency bands, each of the eight spatial clusters, and groups (the four-way interaction of group x frequency bands x spatial clusters x link lengths). Spatial coherence are shown using dashed lines for controls and solid lines for the migraine patients. Asterisks show the significant group differences for each link length (on the x-axis) and each frequency band (colors of asterisks are matched with the frequency bands), based on Bonferroni post-hoc test (M= migraineurs, C=controls).

### Footnotes

1 The reported confidence internals in this paper are within standard mean error (SE).

